# SARS-CoV-2 antigen exposure history shapes phenotypes and specificity of memory CD8 T cells

**DOI:** 10.1101/2021.07.12.21260227

**Authors:** Anastasia A. Minervina, Mikhail V. Pogorelyy, Allison M. Kirk, Jeremy Chase Crawford, E. Kaitlynn Allen, Ching-Heng Chou, Robert C. Mettelman, Kim J. Allison, Chun-Yang Lin, David C. Brice, Xun Zhu, Kasi Vegesana, Gang Wu, Sanchit Trivedi, Pratibha Kottapalli, Daniel Darnell, Suzanne McNeely, Scott R. Olsen, Stacey Schultz-Cherry, Jeremie H. Estepp, the SJTRC Study Team, Maureen A. McGargill, Joshua Wolf, Paul G. Thomas

## Abstract

Although mRNA vaccine efficacy against severe COVID-19 remains high, variant emergence and breakthrough infections have changed vaccine policy to include booster immunizations. However, the effect of diverse and repeated antigen exposures on SARS-CoV-2 memory T cells is poorly understood. Here, we utilize DNA-barcoded MHC-multimers combined with scRNAseq and scTCRseq to capture the *ex vivo* profile of SARS-CoV-2-responsive T cells within a cohort of individuals with one, two, or three antigen exposures, including vaccination, primary infection, and breakthrough infection. We found that the order of exposure determined the relative distribution between spike- and non-spike-specific responses, with vaccination after infection leading to further expansion of spike-specific T cells and differentiation to a CCR7-CD45RA+ effector phenotype. In contrast, individuals experiencing a breakthrough infection mount vigorous non-spike-specific responses. In-depth analysis of over 4,000 epitope-specific T cell receptor sequences demonstrates that all types of exposures elicit diverse repertoires characterized by shared, dominant TCR motifs, with no evidence for repertoire narrowing from repeated exposure. Our findings suggest that breakthrough infections diversify the T cell memory repertoire and that current vaccination protocols continue to expand and differentiate spike-specific memory responses.

---

The continued evolution of SARS-CoV-2 into diverse lineages has led to reduced efficacy of neutralizing antibody responses raised against ancestral strains, including those used in all approved vaccine formulations. Individuals receiving two doses of mRNA vaccine BNT162b2 experienced a dramatic loss in neutralization titers against the Omicron variant^1^. While current protection studies have focused on antibody responses as the key effector mechanism that limits infection, CD8 T cells are likely to play critical roles in the prevention of severe disease^2–6^. Indeed, there are case reports of patients with impaired humoral immunity where efficient T cell responses appear sufficient for viral clearance^7,8^.

In response to the changing landscape of viral evolution and spread, vaccine recommendations have been continually updated to include a booster dose, representing a third immunization at least six months after the initial dose of the Pfizer/BioNTech or Moderna mRNA vaccines. Despite these measures, significant numbers of so-called “breakthrough” COVID-19 cases are being recorded, with individuals becoming infected after two or three vaccine doses or even after prior infection. In all of these settings, adaptive immunity is repeatedly exposed to SARS-CoV-2 antigens, and the effects of this recurrent boosting on the functional profile, magnitude, and specificity distribution of responding T cells remain poorly understood^9,10^. In particular, it is largely unknown if repeated exposure to the same SARS-CoV-2 antigens boosts pre-existing T cell memory and, further, if an exposure to a novel antigen (e.g, infection after vaccination or infection with a new viral variant) induces *de novo* memory and diversifies the TCR repertoire, or instead preferentially expands previously primed responses.

CD8 T cells recognize antigen presented on the cell surface by the Class I Major Histocompatibility Complex (MHC), which is encoded by the most polymorphic genes in the human population (Human Leukocyte Antigen, HLA genes)^11^. Variability of peptide-MHC across and within donors makes measuring epitope-specific T cell responses challenging, and as a result, studies often rely on bulk response assays (e.g., peptide stimulation). Although peptide stimulation assays in principle can provide an estimate of the total magnitude of the CD8 response, they underestimate the frequency of epitope-specific T cells^12^ Further, because these assays require cellular activation to detect a response, they prevent the direct assessment of cell phenotypes *ex vivo*. Staining with MHC-multimers loaded with individual peptides is an alternative approach, which requires pre-selection of immunogenic peptides. Several SARS-CoV-2 epitopes presented by common HLA alleles were discovered in the past two years, permitting the tracking of epitope-specific T cell responses in infected^13–26^ and vaccinated individuals^9,12^ using MHC-multimers.

Here we utilized DNA-barcoded MHC-dextramers with subsequent scRNAseq and scTCRseq to investigate the effects of repeated antigen exposures (SARS-CoV-2 infections and vaccinations with Pfizer/Biontech BNT162b2) on the key features of the CD8 T cell response, including response magnitude, functional gene expression profiles (assessed directly *ex vivo*), and the constituent T cell receptor repertoire. In other contexts, persistent exposure to antigen has been shown to drive various forms of T cell dysfunction, including exhaustion^27^. Further, the focused priming on SARS-CoV-2 spike antigens, the only component of all approved vaccines, may bias subsequent responses during a breakthrough infection towards recall to spike. Thus, it is crucial to understand how pre-existing T cell memory impacts the immune response and memory formation to novel SARS-CoV-2 antigens after repeated exposures.

## Results

### Antibody responses to SARS-CoV-2 infection and vaccination

To investigate the effect of repeated SARS-CoV-2 antigen exposure on pre-existing memory T cells, we selected a cohort of 55 individuals from SJTRC, a prospective, longitudinal study of St. Jude Children’s Research Hospital adult (≥18 years old) employees (Fig. 1a). Sixteen of these participants remained negative for SARS-CoV-2 during weekly PCR testing (naive, N1-N16), whereas 30 of the subjects were diagnosed as SARS-CoV-2 positive with a PCR test and recovered from mild disease (recovered, R1-R30) during the study period. Both the naive and recovered groups received two doses of the Pfizer-BioNTech BNT162b2 mRNA vaccine, and plasma and PBMC samples were collected for all subjects after the second dose of vaccine and at various earlier time points. This produced four subgroups with distinct antigen exposure combinations: infection only (inf, R1-R16), vaccinated only (vax2, N1-N16), infected followed by one dose of vaccine (inf-vax1, R17-R26), and infected followed by two doses of vaccine (inf-vax2, R1-R26). All inf and inf-vax1 subjects were also sampled after their second dose of vaccine, and therefore have matched samples in the inf-vax2 group (Fig. 1b). Additionally, we collected samples from 9 donors who tested positive for SARS-CoV-2 after receiving both doses of BNT162b2 and experienced symptomatic breakthrough infection (vax2-inf, or “breakthrough” group, B1-B9). As expected, the only group negative for N-protein specific antibodies was the vax2 group that was not infected with SARS-CoV-2 (Extended data Fig. 1a). In concordance with previous reports^28–30^, we observed anti-RBD (Fig. 1c, Extended data Fig. 1b) and anti-spike protein IgG (Extended data Fig. 1c) boost after vaccination of recovered individuals. Also in line with other studies^28–33^, most of the antibody boost in SARS-CoV-2 recovered individuals is caused by the first rather than the second vaccine dose, as only two donors (R20, R26) showed a boost in anti-RBD antibody levels after the second vaccine dose, while antibody levels in other donors remained stable (Extended data Fig. 1b). Overall, anti-RBD (Fig. 1c) and anti-spike IgG levels (Extended data Fig. 1c) were similar between vax2 and inf-vax groups. However, breakthrough cases exhibited significantly, but not dramatically, lower anti-RBD and anti-spike antibody levels after infection compared to both vax2 and inf-vax2 individuals (Fig. 1c).

**Figure 1.**
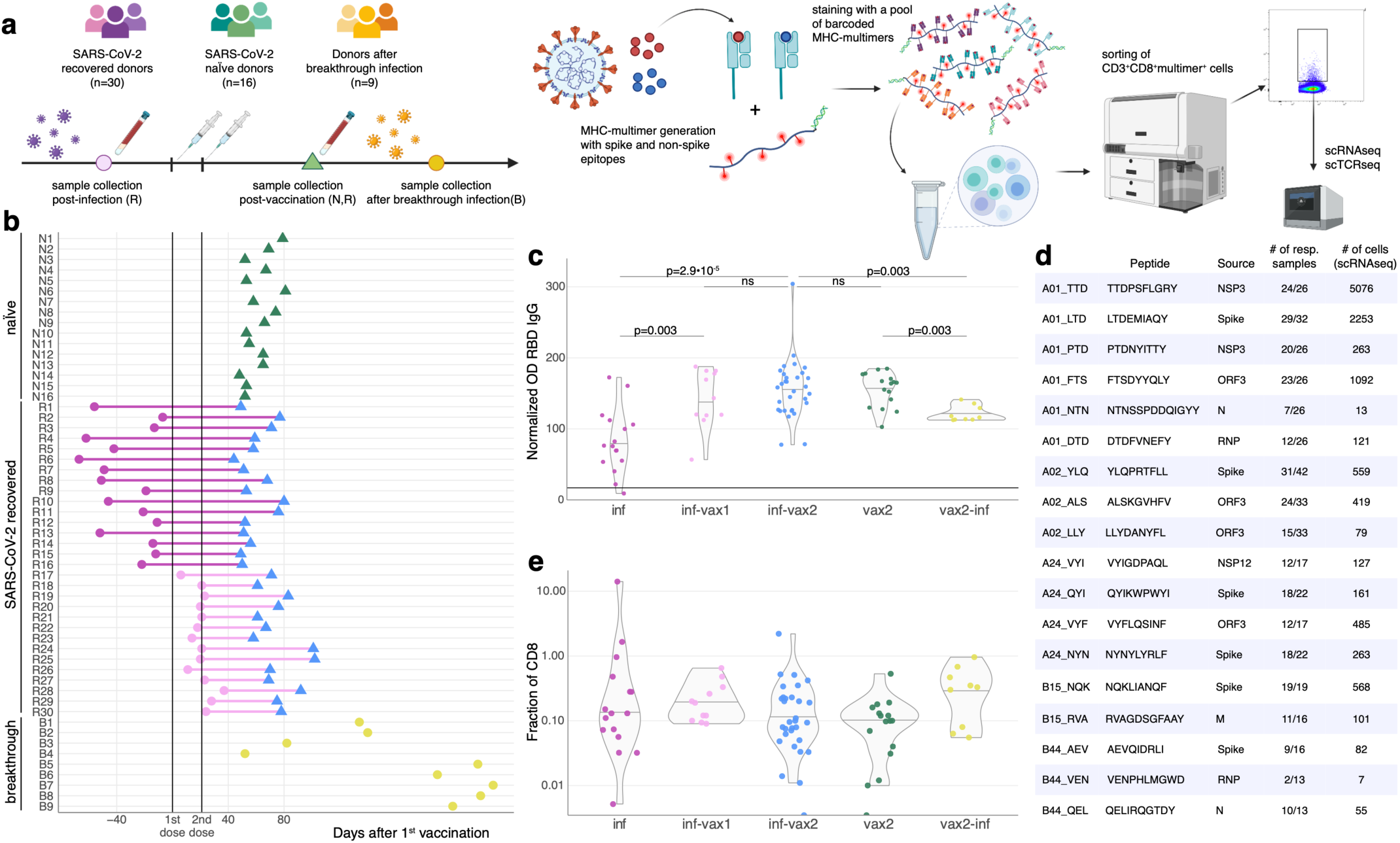
Measuring CD8^+^ T cell epitope-specific responses after diverse SARS-CoV-2 exposures. **a. Study design.** Selected spike and non-spike SARS-CoV-2 T cell epitopes were loaded on recombinant biotinylated MHC-monomers. Resulting peptide-MHC complexes were polymerized using fluorescently labeled and DNA-barcoded dextran backbones. Next, we stained PBMC samples with pools of MHC-multimers, isolated bound cells using FACS, and performed scRNAseq, scTCRseq, and CITEseq using the 10X Genomics platform. **b.** Time of blood sampling for each donor is shown relative to the first dose of mRNA vaccine. **c. Anti-RBD IgG antibody levels in previously infected individuals increase after BNT162b2 vaccination.** Anti-RBD IgG levels in the plasma were determined by ELISA. The normalized OD is the percent ratio of the sample OD to the OD of the positive control for each plate. Plasma was collected from previously infected donors prior (purple, inf), after 1 vaccine dose (inf-vax1, pink), and after 2 vaccine doses (inf-vax2, blue); SARS-CoV-2 naive donors after the full vaccination (vax2, green), and donors that were infected after vaccination (breakthrough, vax2-inf, yellow). All comparisons were done with Mann-Whitney U test, p-values are reported after Benjamini-Hochberg correction. Central line on violin plots depicts the median. **d. List of SARS-CoV-2 epitopes used in this study and summary statistics for resulting epitope-specific response. e. Total frequency of MHC-dextramer-positive cells is similar in all studied groups** (p>0.05 for all pairwise comparisons, Mann-Whitney U test after multiple test correction). Percentage of MHC-multimer-positive cells from all CD8^+^ T cells measured by flow cytometry is shown on a log_10_-scale. Central line on violin plots shows the median.

### Magnitude of epitope-specific CD8^+^ T cell response to mRNA infection and vaccination

To evaluate epitope-specific CD8^+^ T cell responses to SARS-CoV-2 antigen exposure, we investigated previously published data for spike-derived epitopes with a resolved HLA-restriction confirmed in multiple publications. This search resulted in the selection of six spike protein epitopes presented on the HLA alleles A*01:01, A*02:01, A*24:02, B*15:01, and B*44:02^13,15,17,20,24–26,34–36^. We then added 12 previously described non-spike epitopes presented on the same HLA molecules, resulting in a total panel of 18 SARS-CoV-2 epitopes (Fig. 1d, Extended data Table 1). In addition, four of the selected epitopes (A24_VYI, B15_NQK, B44_AEV and B44_VEN) were highly similar to orthologs from common cold coronaviruses (CCCoV), and the CCCoV variant pMHC-dextramers were also included to test the cross-reactive potential of these epitopes^37–40^.

PBMCs from each donor were stained with a panel of DNA-barcoded, fluorescently labeled dextramers (Fig. 1a, Extended data Table 1) that matched the donor’s HLA alleles (Extended data Table 2). For vax2 donors, these panels only included spike-derived dextramers. Epitope-specific T cells (CD3^+^CD8^+^dextramer^+^ cells) were isolated using FACS (Extended data Fig. 2) and then assayed with scRNAseq, scTCRseq, and CITEseq using the 10x Chromium platform. We observed a detectable (>0.01%) dextramer-positive CD8^+^ T cell response in 15/16 vaccinated donors that were not previously infected and in 37/39 SARS-CoV-2-infected donors. Although the overall frequency of dextramer-specific cells was low (0.41±0.17% SEM of CD8^+^ T cells; range: 0.01-14.1% of CD8^+^ T cells), it was comparable to the epitope-specific memory cell frequencies observed months after challenge in other studies of SARS-CoV-2 infection^13,17,20,21^, even though these studies frequently used peptide stimulation covering an entire protein or multiple proteins. Furthermore, the absolute magnitude of the epitope-specific T cell responses was similar across all groups (Fig. 1e) despite varying sources (vaccine/infection) of antigen exposure (p>0.05 for all pairwise comparisons, Mann-Whitney U test with Benjamini-Hochberg multiple testing correction).

### HLA-B*15:01 presents a spike-derived epitope cross-reactive to CCCoV

Use of the DNA-barcoded dextramers allowed us to deconvolve the overall T cell response to 18 distinct epitope-specific responses. For each cell, we calculated the number of unique molecular identifiers (UMIs) per dextramer, and considered a cell as dextramer-specific if more than 30% of the dextramer-derived UMIs corresponded to that dextramer’s specific barcode. Cells that did not match the criteria (i.e., exhibited ambiguous binding or fewer than 4 UMIs per most abundant dextramer) were considered unspecific binders and were excluded from the dataset. This resulted in non-overlapping dextramer-positive and -negative groups of cells for each dextramer (Fig. 2a, Extended data Fig. 3). To further assess this threshold, we considered the dextramer assignment of individual cells among the 43 most abundant T cell clones (i.e., clonotypes with ≥ 20 cells) as defined by scTCRseq. Of these clonotypes, 72% (31/43) matched a single epitope across all cells (Fig. 2b), with only six of the most abundant clonotypes assigned to several non-orthologous epitopes. However, for all these clonotypes, there was a clear dominant epitope assigned to the majority of cells, demonstrating the general robustness of the dextramer specificity thresholds. Interestingly, five of the most abundant TCR clonotypes were assigned to both B15-NQK_Q SARS-CoV-2 and B15-NQK_A CCCoV (HKU1/OC43) orthologs of the spike epitope, supporting our initial hypothesis for potential SARS-CoV-2/CCCoV epitope cross-reactivity. Indeed, the UMI counts for the dextramers with SARS-CoV-2 and CCCoV variants of the epitope were strongly correlated (Fig. 2c), suggesting that the exact same cells can bind both versions of the epitope.

**Figure 2.**
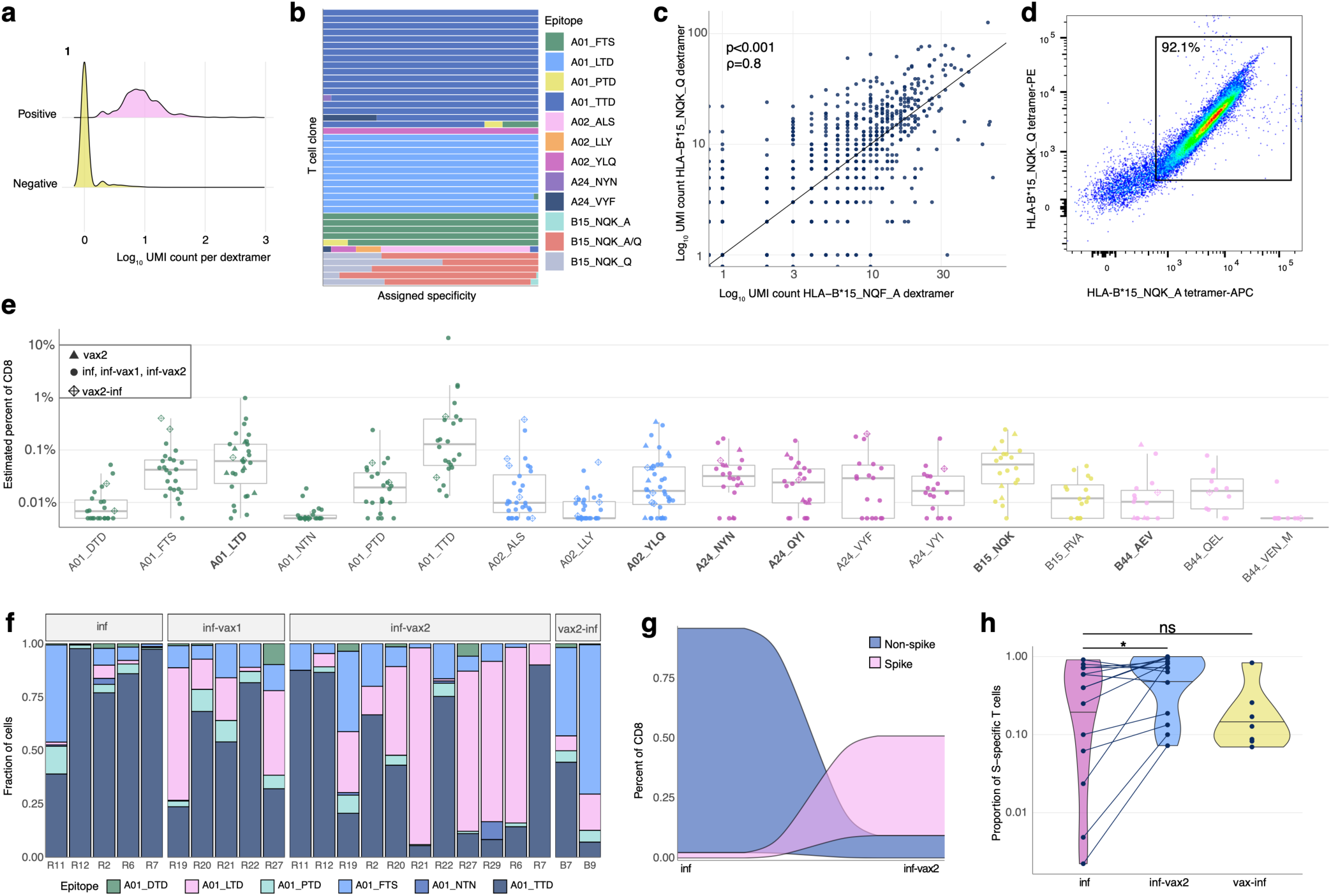
Magnitude, dynamics, and cross-reactivity of CD8^+^ epitope-specific responses after diverse SARS-CoV-2 exposures. **a**. **Antigen specificity of each T cell inferred from dextramer-barcode UMI counts.** Representative distribution of the number of UMIs in cells called dextramer-positive (pink) and dextramer-negative (yellow). **b. T cells within a clone have largely consistent specificity assignments, except T cells that cross-react with common cold coronavirus epitopes (B15_NQK_A/B15_NQK_Q pair).** Each bar shows a fraction of cells of a given clonotype attributed to different dextramers. The 43 most abundant clones (more than 20 cells) are shown. **c. The correlation between the number of UMIs for B15_NQK_Q (SARS-CoV-2) and B15_NQK_A (OC43 and HKU1) dextramers (Spearman ρ=0.8, p<0.001)**. **d. Cross-reactivity between HLA-B*15:01-NQK epitope variants confirmed *in vitro*.** Jurkat cell line expressing αβTCR identified from scTCRseq data binds pMHC multimers loaded with both SARS-CoV-2 and CCCoV variants of the epitope. **e. The magnitude of epitope-specific CD8^+^ T cell responses.** Each point depicts an estimated frequency of epitope-specific T cells in a sample. Estimated frequency was calculated as a fraction of dextramer-specific T cells in scRNAseq results multiplied by bulk frequency of dextramer-stained CD8^+^ cells of all CD8^+^ cells measured by flow cytometry. Central line on boxplot shows the median. Epitopes from spike protein are in bold font. **f. Composition of HLA-A*01-restricted T cell response in HLA-A*01 positive donors.** Increasing proportion of spike-targeting T cells (pink) is observed after vaccination of infected individuals. **g. Boosting of spike-specific epitope fraction after vaccination (donor R6). h. Previously infected individuals have a higher proportion of spike-specific T cells after vaccination than before vaccination** (p=0.025, one-sided Wilcoxon signed-rank test). Spike T cell proportion (shown on a log_10_-scale) was calculated as a fraction of spike-specific T cells out of all CD8^+^ epitope-specific T cells of a donor in scRNAseq data. Central line on the violin shows the median.

To further validate that a single TCR could recognize both variants of B15-NQK, we made a Jurkat cell line expressing one of the potentially cross-reactive αβTCRs. This T cell line recognized both CCCoV and SARS-CoV-2 variants of the peptide, as demonstrated by HLA-B*15:01-multimer staining (Fig. 2d) and peptide stimulation assays (Extended data Fig. 4). Interestingly, the presence of T cells specific to this epitope coincided with higher IgG levels against the spike protein of common cold betacoronaviruses HKU1 and OC43 prior to infection or vaccination (Extended data Fig. 5). These data indicate that SARS-CoV-2 may reactivate cross-reactive memory CD8^+^ T cells established during previous OC43/HKU1 infections.

### Spike vs. non-spike response distribution varies with antigen exposures

Because barcoded dextramers allow us to simultaneously measure the response to multiple epitopes in the same sample at single-cell resolution, we also utilized these data to compare the magnitude of the response to different epitopes. Among all the tested epitopes, A01_TTD, A01_LTD, A02_YLQ, and B15_NQK elicited the strongest overall response (Fig. 2e) and were also found in the majority of HLA-matched samples. Although we observed responses to all other epitopes, they occurred at lower frequencies and only in a subset of HLA-matched donors.

Donors with distinct HLA alleles present different subsets of epitopes. Thus, to robustly compare the magnitude of spike and non-spike responses, we characterized the contribution of each of the six A*01:01 restricted epitopes in HLA-A*01:01-positive SARS-CoV-2 convalescent individuals (n=13). Interestingly, the proportion of the spike-derived epitope A01_LTD response significantly increased in inf-vax2 individuals compared to infected individuals prior to vaccination (0.8% A01_LTD-specific cells of total A01-restricted response for inf-only group, vs 48% A01_LTD-specific cells of total A01-restricted response for inf-vax2, p<0.0001 Fisher exact test; Fig. 2f). Similar but less striking effects were also observed within HLA-A*02:01-positive individuals (n=19) for three A*02:01 restricted epitopes (33% of A02_YLQ-specific cells for inf-only, vs 82% of A02_YLQ-specific cells for inf-vax2, p<0.0001 Fisher exact test; Extended data Fig. 6). These patterns suggest that the distribution of T cell specificities was shifted towards spike-derived epitopes following vaccination of these previously infected donors (Fig. 2g, Extended data Fig. 7). Indeed, among all donors regardless of HLA type, we observed a significant increase in the fraction of the spike-specific T cell response after vaccination, indicating the recall of epitope-specific memory T cells among previously infected individuals as a result of vaccination (Fig. 2H, p=0.025, one-sided Wilcoxon signed-rank test). Similar to the antibody response, most of this expansion was likely due to the first rather than second dose of the vaccine, as we did not observe a T cell boost between the first and second doses of vaccine in 7/10 subjects (Extended data Fig. 8). In sum, vaccination is able to potently and selectively expand spike-specific responses.

Given the potent induction and expansion of spike-specific responses by vaccination, even in individuals who were previously infected, we predicted that infection of previously vaccinated individuals (breakthrough, vax2-inf) would maintain a spike-specific bias. Surprisingly, we observed a large non-spike-specific T cell response in the majority of the breakthrough 6/7 (vax2-inf) cohort (Fig. 2h), indicating that a robust primary response to non-spike SARS-CoV-2 antigens during the breakthrough infection is not impaired by the presence of spike-specific immune memory elicited by vaccination. The ratio between spike-and non-spike-specific T cells in breakthrough cases (vax2-inf) was no different from that of donors who were only infected (inf; p=0.97, Mann-Whitney U test), indicating that the T cell response to the non-spike antigens is of comparable magnitude among those who were only infected and and those who experienced breakthrough infection after vaccination (vax2-inf). Thus, while the magnitude of the epitope-specific responses is similar across all exposure types, the composition of epitope-specific responses is clearly skewed by both the number and order of exposures.

### Phenotypes of epitope-specific CD8^+^ T cells following SARS-CoV-2 infection and vaccination

To understand if different types of antigen exposures could also drive divergent phenotypes among epitope-specific T cells, we leveraged the single-cell gene expression (scGEX) data corresponding to our TCR and dextramer data. Unsupervised clustering identified 11 distinct transcriptional subsets of epitope-specific cells (Fig. 3a). These clusters were manually annotated using the surface abundance of conventional memory markers (CCR7 and CD45RA) measured by CITEseq (Fig. 3b) and other well-studied expression markers (Fig 3c, Extended data Table 3, Table 4), allowing us to identify the following populations: Transitional Memory (Effector memory(EM)/EM with re-expression of CD45RA(EMRA)), EMRA-like, Central Memory (CM)/T stem cell-like memory (Tscm), Differentiated effectors, naive/Tscm, EM, Resting effectors, EM with exhaustion markers, Resting memory, CM with *GATA3*, and Cycling. Though the proportions of these T cell populations varied substantially across antigen exposure contexts, each gene expression cluster contained cells from all five exposure groups (Fig. 3d, Extended data Fig. 9, 10, 11). Natural infection, breakthrough cases, and vaccination led to the formation of potent T cell memory, including highly cytotoxic populations (clusters 0,1,3,5) and populations with expression of common markers of durable cellular memory (clusters 2,4,8,9), e.g. *TCF7, IL7R,* and *CCR7* (Fig. 3c).

**Figure 3.**
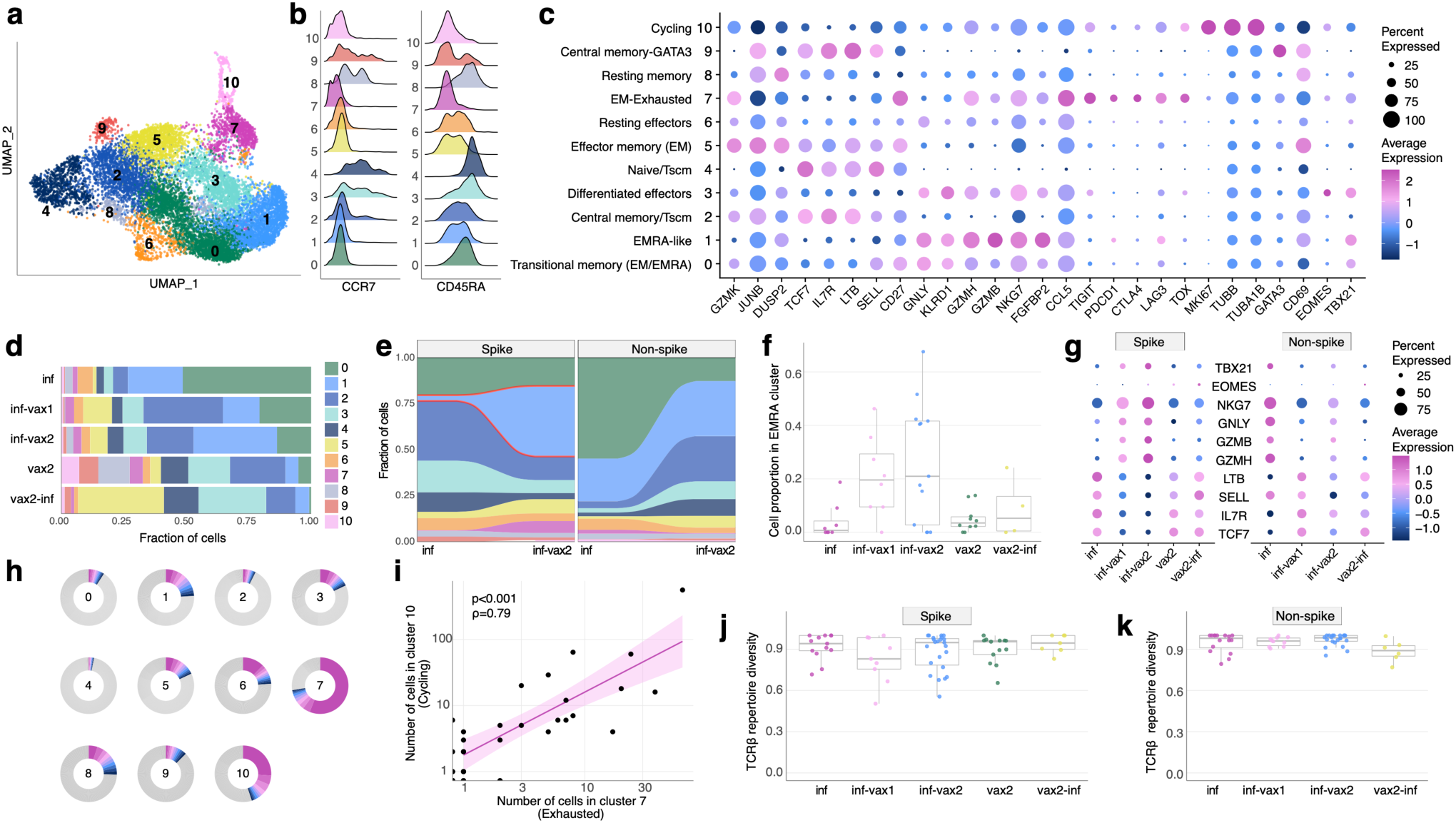
Phenotypic diversity of epitope-specific CD8^+^ T cells after diverse SARS-CoV-2 exposures. **a. UMAP (Uniform manifold approximation and projection) of all SARS-CoV-2 epitope-specific CD8 T cells based on gene expression (GEX)**. Color shows results of graph-based unsupervised clustering performed with the Seurat package. **b. Density plot of CCR7 and CD45RA surface expression (measured by CITE-seq) in GEX clusters. c. Bubble plot of representative differentially expressed genes for each cluster**. Size of the circle shows percentage of cells in a cluster expressing a certain gene, color scale shows gene expression level. **d. Distribution of epitope-specific T cells in gene expression clusters between study groups. e. Proportion of spike-specific T cells is significantly increased in cluster 1 after vaccination of previously infected individuals, compared to the pre-vaccination timepoint (p<0.0001, Fisher exact test). f. Proportion of spike-specific cells in EMRA (cluster 1) across study groups for samples with more than ten spike-specific cells (Kruskal-Wallis H test p=0.028)**. Central line on boxplot shows the median. **g. Expression of classical cytotoxic and memory markers across study groups and T cell specificities**. Size of the circle shows percentage of cells in a cluster expressing a certain gene, color scale shows gene expression level. **h. Clone size distribution within GEX clusters**. Fractions of cells from 10 most abundant clonotypes in each cluster are shown with colors, all other clonotypes are shown in grey. **i. Number of cells in cluster 7 (Exhausted) and cluster 10 (Cycling) in samples are strongly correlated (Spearman ρ=0.79, p<0.001)**. Shaded area shows 95% confidence interval for linear fit. **j-k. T cell repertoire diversity of spike (j) and non-spike specific repertoires across study groups** (p=0.63 for spike, p=0.17 for non-spike, Kruskal-Wallis H test). Normalized Shannon entropy of TCRβ is plotted for samples with more than 3 unique TCRβ clonotypes. Central line on boxplot shows the median.

### Repeated exposures cause a shift of T cell memory phenotypes towards EMRA

To determine if a vaccine-induced recall response affects the phenotypes of T cells, we compared the GEX cluster distribution between inf-only and inf-vax2 donors. We observed a significant post-vaccination shift towards a more highly differentiated effector phenotype (EMRA, cluster 1) of spike-specific cells (Fig. 3e, S10, p<0.0001, Fisher exact test). Interestingly, there was no such change for non-spike-specific cells, suggesting that vaccination specifically increased the proportion of cells in cluster 1 (EMRA-like) among SARS-CoV-2 recovered donors via a recall of spike-specific memory T cells (Fig. 3f). Indeed, inf-vax1 and inf-vax2 groups were characterized by spike-specific T cells with higher *GZMB, GZMH, GNLY,* and *NKG7* expression and lower *TCF7, IL7R, SELL,* and *LTB* expression than those in other groups, consistent with the EMRA phenotype (Fig. 3g). Interestingly, the spike-specific T cells in breakthrough infections (vax2-inf) exhibited expression profiles more similar to groups with a single type of antigen exposure (vax2 or inf) than to those of inf-vax1,2 subjects.

### Repeated SARS-CoV-2 antigen exposure does not lead to an exhausted T cell phenotype

Repeated or chronic antigen exposure leads to T cell exhaustion in multiple experimental models^27^. Several publications have linked T cell exhaustion to an impaired SARS-CoV-2 cellular response^41–43^. While our epitope-specific data similarly included a cluster with high expression of classical exhaustion markers (cluster 7, EM-Ex, Fig. 3c), including *CTLA-4, PD-1, TOX,* and *TIGIT*, this cluster was present in multiple donors (26/51) across all groups, including vax2-only (Extended data Fig. S9, S10). In concordance with previous reports^23,42^, this “exhausted cluster” was extremely clonal in composition (Extended data Fig. 12), with more than 70% of the cluster repertoire occupied by just 10 clones (Fig. 3h). We also observed that the number of cells in the “exhausted cluster” within a patient strongly correlated with the number of cells in the cluster of cycling cells (Fig. 3i). Thus, the presence of the exhausted cluster is connected to both clonal expansion and cell proliferation, suggesting that donors who have such cells are still in the active rather than memory state of the CD8 T cell response. To test this, we looked at the distribution of cells among clusters at two available time points after infection (Fig. 1b, donors R1-R30, average time between timepoints was 75.5 days, range 40-126). The number of cells in cluster 7 declined with time (Extended data Fig. 13), indicating that this “exhausted” subset is both common among mild infections yet transient and, importantly, that the presence of these cells is not sufficient to cause notable pathology. Rather, the exhaustion phenotype appears primarily correlated with time since antigen exposure.

### Convergent and diverse TCR repertoire of epitope-specific CD8^+^ cells

Our data thus far indicate that vaccination after infection boosts pre-existing T cell memory to spike antigens and leads to significant alterations in the cellular phenotypes. We next asked whether this recall response affects the diversity of the underlying recruited T cell receptor repertoires, potentially narrowing repertoire diversity after each exposure. To compare the TCR repertoires of epitope-specific cells elicited in response to different exposure contexts, we assessed the overall TCRβ repertoire diversity (represented by normalised Shannon entropy). The diversity of both spike- (Fig. 3j) and non-spike-specific repertoires (Fig. 3k) was comparable among all groups (p=0.63 for spike, p=0.17 for non-spike, Kruskal-Wallis H test), suggesting that a diverse repertoire of T cells persists in the memory compartment regardless of antigenic history and is not narrowed by the recall response. This is especially notable among the breakthrough infections, as it indicates that these individuals mount *de novo* diverse non-spike-specific T cell memory in response to the infection.

We and others have previously shown that T cells recognizing the same epitopes frequently have highly similar T cell receptor sequences^44–46^. We therefore constructed a similarity network of paired, unique αβTCR sequences from our data (Extended data, Table 5), using a threshold on the TCRdist^44^ similarity measure to identify highly similar clonotypes (Fig. 4a). The clusters of similar sequences almost exclusively consisted of TCRs with the same epitope specificity and feature biases in V-segment usage (Extended data Fig. 14, 15, 16), as well as striking positional enrichment of certain amino acid residues within the CDR3 region (Fig. 4b). We next individually cloned 12 of these TCRs from the 7 largest similarity clusters into a TCR-null Jurkat cell line (Extended data, Table 6). The resulting cell lines exhibited the expected specificity based on dextramer barcodes both in peptide stimulation assays (Extended data Fig. 17) and dextramer staining (Extended data Fig. 18), validating both the bioinformatics approach and the reagents. We next asked if the same motifs were recruited into the response across antigen exposure histories. Importantly, many of the confirmed CDR3 motifs from spike-specific TCRs were shared among donors who recovered from natural infection, including breakthrough infections, and among immunologically naive donors after vaccination (Fig. 4c). This suggests that epitope recognition is achieved by the same TCR-pMHC molecular interactions regardless of the method of antigen exposure, and thus one could expect similar specificity to potential epitope variants for memory T cells elicited by vaccination or natural infection.

**Figure 4.**
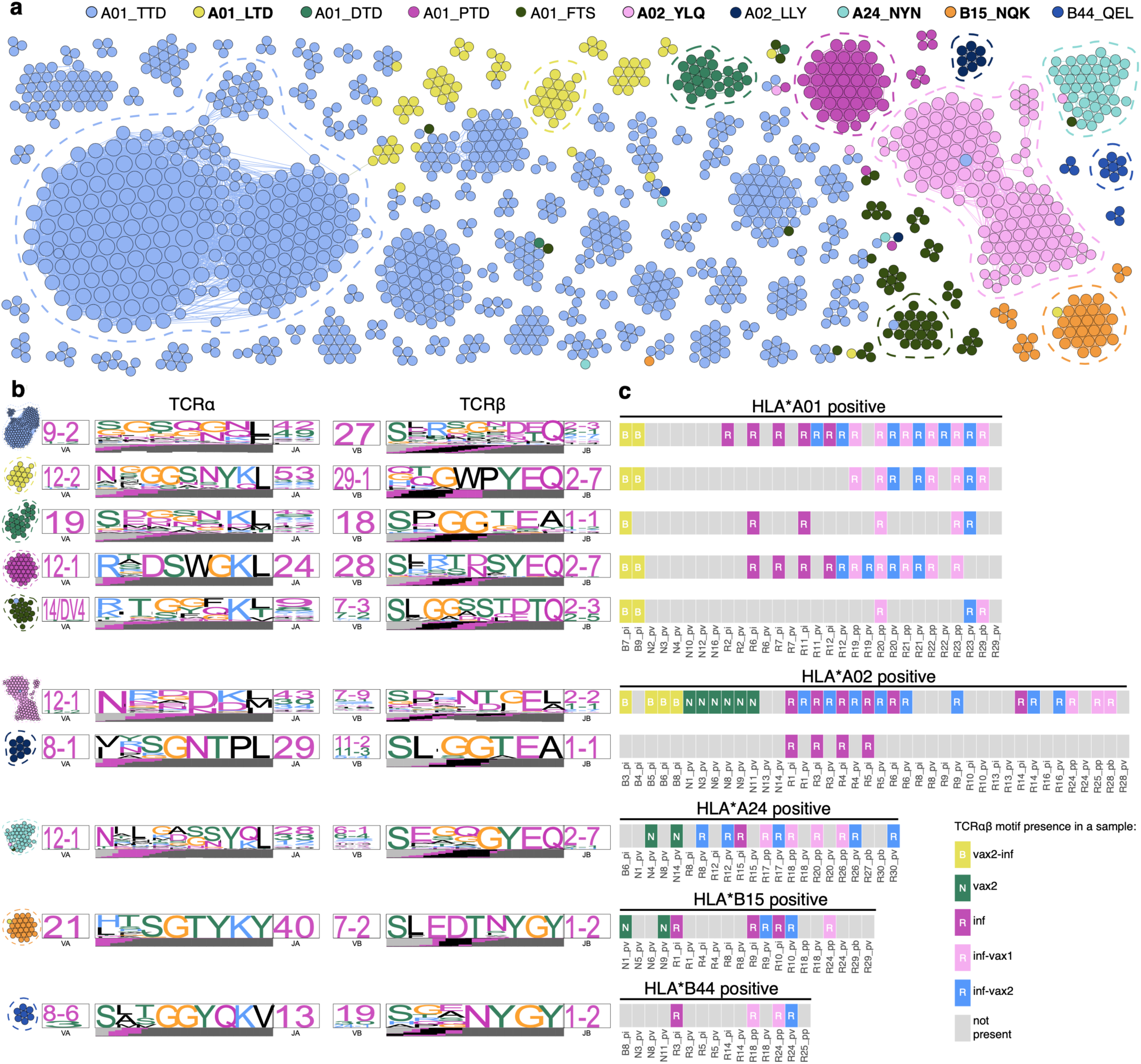
Diverse polyclonal repertoires of epitope-specific T cells after diverse SARS-CoV-2 exposures. **a. SARS-CoV-2 epitope-specific αβTCR amino acid clonotypes feature clusters of highly similar sequences with the same epitope specificity**. Each node on a similarity network is a unique paired αβTCR amino acid sequence, and an edge connects αβTCRs with TCRdist less than 110. Each color represents a certain epitope specificity. Only clusters with more than two members are shown. Spike-derived epitopes are in bold font. **b. TCR amino acid sequence motifs of α and β chains (TCRdist logos) for the largest clusters of highly similar TCRs for each epitope (circled with dashed line on a). c. TCRs with the same sequence motifs are found across all study groups in a matching HLA-background. Occurrence of TCR motifs on the left is shown for all HLA matching samples (rectangles on the plot)**. Grey rectangles represent samples lacking the TCR motif. The color of the rectangle that has a TCR motif corresponds to the sample group.

### TCR motifs recognize most mutated epitopes in SARS-CoV-2 variants

Our TCR analyses established that regardless of antigen history, the same dominant TCR motifs were utilized by subjects responding to a number of important SARS-CoV-2 epitopes. Thus, these TCRs can be used to probe how memory responses from these exposures will detect epitopes in variant SARS-CoV-2 strains. To investigate the potential impact of SARS-CoV-2 variants on T cell recognition, we searched the GISAID for mutations in the selected CD8 epitopes. Mutations in both current and previous viral lineages were included in the analysis if they appeared in at least 10% of a Pango lineage and in at least ten thousand isolates. Notably, no mutations in the studied epitopes were observed in the Omicron variant. However, we identified 10 mutations among the 200 Pango lineages, including Delta and Gamma WHO variants of concern. Models predicting peptide-MHC binding (NetMHCpan4.1b^47^) suggest that these mutations do not impact the binding of the epitope to the restricting HLA allele, as both mutated and wild-type epitope variants are predicted to be strong binders (Extended data Table 7). Thus, we decided to test whether our transgenic TCR lines were capable of recognizing these mutated epitopes. All three mutated epitopes of A01_TTD could be recognized by at least one of our A01_TTD-specific T cell lines. (Extended data, Fig. 19). Interestingly, one of the mutated A01_TTD epitopes (TT***N***PSFLGRY) was recognized by one of the two generated TCR lines, highlighting the importance of TCR diversity in the cross-reactivity to novel variants. Neither A02_YLQ-specific TCR line was activated by the mutant S:P272L epitope YLQPRTFLL (Extended data, Fig. 19), confirming the data from Dolton et al. This mutation was speculated to play a role in a second Europe COVID-19 wave in summer-autumn of 2020^48^. However, none of the currently abundant SARS-CoV-2 variants bear this or any other variant of the A02_YLQ epitope at large frequency. Of the four mutations observed in the A24_NYN epitope, two escaped recognition by both cloned TCR lines (Extended data, Fig. 19). The mutation L452R affecting A24_NYN is of particular interest as it is present in over 95% of all Delta variant sequences in GISAID. Whether individuals infected by the Delta variant could utilize other TCR motifs to recognise this mutated epitope requires further investigation. Together, our data suggest that the T cell memory repertoire established by SARS-CoV-2 infection or vaccination has great cross-reactivity potential against novel viral variants, and further shows that not all of the viral mutations affecting T cell epitopes result in the T cell immune escape, even from the most public TCR clones.

## Discussion

Understanding the effects of multiple antigen exposures, in various contexts, on the development of effective CD8^+^ T cell memory against SARS-CoV-2 is important for determining susceptibility to subsequent infections and the potential for booster vaccination to improve outcomes. To address this, we analyzed multiple parameters of the CD8^+^ T cell response across five types of antigen exposure history and found that repeated antigen exposures (up to three) continued to induce expansion to the included antigens and drive further functional maturation. Despite this, the underlying TCR repertoire structure within epitope specific responses maintained diversity, which is a promising indication of continued vaccine efficacy. Narrowing of TCR diversity has been shown in a number of contexts to correlate with poor immunological control. As fourth boosters and increased rates of breakthrough infections are providing additional exposures, these data are a useful benchmark for determining how these relatively rapid repeat exposures will continue to mature the response. Close monitoring of these important parameters—magnitude, functional profile, and repertoire diversity—should be continued in longitudinal cohorts with diverse antigen exposures.

Breakthrough infections of vaccinated individuals have a much lower risk of causing severe disease but are a concern for maintaining transmission and exposing vulnerable populations. Furthermore, breakthrough infections have increased with greater serological drift in emerging variants of concern, including Delta and Omicron. We found that functional profiles among breakthrough infections (vax2-inf) were distinct from other forms of antigen exposure but consistent with effector T cell differentiation and, in fact, demonstrated an arguably earlier differentiation state than inf-vax2 individuals. In addition, we show that these individuals form non-spike specific T cell memory at robust levels, indicating that there is not an intrinsic defect among these individuals in mounting robust anti-SARS-CoV-2 responses and diversifying the T cell memory pool to SARS-CoV-2 internal proteins. The proportion of breakthrough subject response targeting spike epitopes was in fact smaller than that of the inf-vax2 subjects, indicating that individuals with breakthrough infection were not preferentially biased towards spike responses. This is especially important given the continued emergence of SARS-CoV-2 variants^10,49–51^ and the current uprise in breakthrough infection rate.

In the midst of characterizing T cell responses against SARS-CoV-2-specific epitopes, we also discovered T cells that are cross-reactive for SARS-CoV-2 and common cold coronavirus variants of an HLA-B*15-restricted immunodominant epitope. The possibility of this cross-reactivity was hypothesized previously^52^, where the clonotypes with this TCR motif were the most expanded in an HLA-B*15 positive donor. An epitope from N-protein HLA-B*07_SPR has also been shown to be cross-reactive with HKU1 and OC43 common cold coronaviruses^14,53^, although other studies of T cells specific to these epitopes concluded they were not cross-reactive^5,18^. The extent of protection in HLA-B*15+ and HLA-B*07+ donors recently infected with common cold coronaviruses is yet to be determined, but a high frequency of cross-reactive CD8 T cells may correlate with protection.

The most striking differences we observed based on antigenic history were in the phenotype of elicited cells. In particular, we found an increase in the fraction of EMRA spike-specific T cells following vaccination in previously infected subjects (inf-vax2). It remains unclear whether the EMRA phenotype is associated with more or less durable and efficient protection, and longer follow-up studies of the durability of memory in vax2-only, inf-only, and inf-vax2 groups should closely monitor the phenotype of antigen-specific T cell responses and their persistence. This is particularly relevant given the current routine practice of third, and soon possibly fourth, vaccine doses.

Precise measurement of epitope-specific T cell and B cell responses is crucial for defining the correlates of SARS-CoV-2 protection, which will inform vaccination strategies to prevent pandemic recurrence as additional SARS-CoV-2 variants emerge. The striking similarity between the magnitude and constituent repertoires of epitope-specific CD8 T cell responses following infection, vaccination, or infection followed by vaccination, indicate that mRNA vaccines are capable of inducing nearly equivalent memory as an infection episode and further expanding previously established responses. These data further suggest that booster shots, if needed to address antibody-escape to Omicron and other variants, will not substantially alter the repertoires of established anti-spike T cell memory.

Our data have also provided a useful confirmation of the specific sequence features of several SARS-CoV-2 epitope-specific responses. The generation of monoclonal T cell lines that can be used to rapidly survey variant peptides provides an analogous tool as a monoclonal antibody for characterizing antibody escape mutations. Here, we were able to show subtle variations in the loss of recognition by multiple TCR lines recognizing the same epitope. These tools can be used to screen emerging variants of concern and also predict mutations that might lead to relevant epitope escape.

Our study has several limitations that should be considered. First, we focus on comparisons between T cells specific for a pre-selected set of CD8^+^ epitopes previously identified in large epitope discovery studies^54^. This set of epitopes, although considerable in size given the nature of our experiments, does not necessarily cover all immunodominant responses, and may also exclude novel epitopes induced only by vaccination (though to date none have been reported). Furthermore, the epitopes chosen are presented on a limited subset of HLA-alleles that, while abundant in populations of European ancestry, are less representative of other populations. Additional epitope discovery studies of SARS-CoV-2 and other clinically relevant pathogens covering more HLA alleles from cohorts of diverse ancestry are important to overcome current biases in the literature and integral for fully elucidating the complex interactions between genotype, phenotype, and environment on the immune response. Secondly, we were only able to analyze a relatively small number of breakthrough infection cases. Our data suggest that, going forward, it will be important to more exhaustively profile the epitope-specific responses of individuals who experience breakthrough infections, particularly by obtaining prospective samples after vaccination but prior to infection.

In addition, we only had access to PBMC samples, which do not allow study of the distinct features of the cellular response at the site of infection. Particularly in breakthrough infections, if differential trafficking of memory cells to the airways occurred, it may bias our interpretation of the observed response. Lastly, the variation in our sampling times across all subgroups may introduce additional noise due to active T cell response dynamics. More regular and frequent sampling in a larger cohort of fully vaccinated individuals will facilitate a more exhaustive understanding of the correlates of protection from SARS-CoV-2 infection and the mechanisms underlying breakthrough infection.

## Online methods

### Human cohort

The St. Jude Tracking of Viral and Host Factors Associated with COVID-19 study (SJTRC, NCT04362995) is a prospective, longitudinal cohort study of St. Jude Children’s Research Hospital adult (≥18 years old) employees. The St. Jude Institutional Review Board approved the study. Participants provided written informed consent prior to enrollment and completed regular questionnaires about demographics, medical history, treatment, and symptoms if positively diagnosed with SARS-CoV-2 by PCR. Study data are collected and managed using REDCap electronic data capture tools hosted at St. Jude^55,56^. Participants were screened for SARS-CoV-2 infection by PCR weekly when on the St. Jude campus. For this study, we selected a cohort of 55 individuals, 16 of which had never tested positive for SARS-CoV-2 (N1-N16), and 39 of which were diagnosed as SARS-CoV-2 positive with a PCR test and recovered from mild disease (R1-R30, breakthrough B1-B9) during the study period. All individuals in this study received two doses of the Pfizer-BioNTech BNT162b2 mRNA vaccine. Vaccination data, including vaccine type and date administered, were obtained from the institutional database which required direct confirmation of vaccine administration records before data entry. Previously infected and naive vaccinated individuals (inf-vax2 and vax2) were sampled at similar time points after their vaccine regimen was complete (R1-R30: 45.5±2.8 SEM, range 25-81 days; N1-N16: 40.7±2.7 SEM, range 23-60 days). Finally, the individuals chosen for each group were of similar ages (R1-R30: 44.2±2.5 SEM, range 23-68 years; N1-N16: 44.1±3 SEM, range 29-73 years; B1-B9: 40.1±4.2 SEM, range 24-60 years). For this study, we utilized the convalescent blood draw for SARS-CoV-2 infected individuals (3-8 weeks post-diagnosis) and the post-vaccination samples (3-8 weeks after completion of the vaccine series). For breakthrough infections, we used the convalescent blood draw. An infection was considered a “breakthrough” if an individual tested positive for SARS-CoV-2 infection by PCR after receiving two doses of the Pfizer-BioNTech BNT162b2 vaccine. Blood samples were collected in 8 mL CPT tubes and separated within 24 hours of collection into cellular and plasma components then aliquoted and frozen for future analysis. Human cohort metadata can be found in Extended data Table 2.

### HLA typing

High quality DNA was extracted from whole blood aliquots from each participant using the Zymo Quick-DNA 96 Plus Kit (Qiagen). DNA was quantified on the Nanodrop (Thermo Scientific). HLA typing of each participant was performed using the AllType NGS 11-Loci Amplification Kit (One Lambda; Lot 013) according to manufacturer’s instructions. Briefly, 50 ng DNA was amplified using AllType NGS 11-Loci amplification primers. The amplified product was then cleaned and quantified on the Qubit 4.0 (Invitrogen). Library preparation of purified amplicons was carried out as described in the protocol, and the AllType NGS Index Flex Kit (Lot 011) was used for barcoding and secondary amplification. Purified, barcoded libraries were quantified using the Qubit DNA HS kit (Invitrogen) and pooled according to the One Lambda Library Pooling table. Pools of up to 48 libraries were then purified and quantified on the TapeStation D5000 (Agilent) before sequencing on a full MiSeq lane at 150×150bp following manufacturer’s sequencing specifications. HLA types were called using the TypeStream Visual Software from One Lambda. HLA typing results can be found in Extended data Table 2.

### Dextramer generation and cell staining

Peptides with >95% purity were ordered from Genscript and diluted in DMSO to 1 mM. pMHC monomers (500 nM) were generated with easYmer HLA class I (A*01:01, A*02:01, A*24:02, B*15:01, B*44:02) kits (Immunaware) according to the manufacturer’s protocol. To generate DNA-barcoded MHC-dextramers we used Klickmer technology (dCODE Klickmer, Immudex). 16.2 µL of HLA monomer (500 nM) were mixed with 2 µL barcoded dCODE-PE-dextramer to achieve an average occupancy of 15 and incubated for at least 1 hour on ice prior to use. Individual dextramer cocktails were prepared immediately before staining. Each cocktail had 1.5 µL of each HLA-compatible barcoded MHC-dextramer-PE and 0.15 µL 100 µM biotin per dextramer pre-mixed to block free binding sites. Samples were divided into 3 batches, and timepoints from the same donor were always processed simultaneously. Donor PBMCs were thawed and resuspended in 50 µL FACS buffer (PBS, 0.5% BSA, 2 mM EDTA). Cells were stained with 5 µL Fc-block (Human TruStain FcX, Biolegend 422302) and a cocktail of dextramers for 15 minutes on ice. After this a cocktail of fluorescently-labeled surface antibodies (2 µL of each: Ghost Dye Violet 510 Viability Dye, Tonbo Biosciences 13-0870-T100; anti-human CD3 FITC-conjugated (Biolegend 300406, clone UCHT1), anti-human CD8 BV711-conjugated (Biolegend, 344734, clone SK1)) and TotalSeq-C antibodies (1 µL anti-human CCR7 (Biolegend 353251), 1 µL anti-human CD45RA (Biolegend 304163)) and 2 µL of TotalSeq-C anti-human Hashtag antibodies 1-10 (Biolegend 394661, 394663, 394665, 394667, 394669, 394671, 394673, 394675, 394677, 394679) were added. Samples were incubated for 30 minutes on ice. Single, Live, CD3-positive, CD8-positive, dextramer-positive cells were sorted into RPMI (Gibco) containing 10% FBS and 1% penicillin/streptomycin using a Sony SY3200 cell sorter. Sorted cells were immediately loaded into a 10x reaction. Chromium Next GEM Single-Cell 5’ kits version 2 (10x Genomics PN: 1000265, 1000286, 1000250, 1000215, 1000252 1000190, 1000080) were used to generate GEX, VDJ and Cite-Seq libraries according to the manufacturer’s protocol. Libraries were sequenced on Illumina NovaSeq at 26×90bp read length.

### Single-cell RNAseq data analysis

Raw data was processed with Cell Ranger version 6.0.0 (10X Genomics). Three batches were subsequently combined using the aggregate function with default parameters. Resulting GEX matrices were analysed with the Seurat R package version 4.0.4 ^57^. Following standard quality control filtering, we discarded low quality cells (nFeatures <200 or >5000, MT% >5%) and eliminated the effects of cell cycle heterogeneity using the CellCycleScoring and ScaleData functions. Next, we identified 2,000 variable gene features. Importantly, we excluded TCR/Ig genes from variable features, so that the gene expression clustering would be unaffected by T cell clonotype distributions. Next, we removed all non-CD8 cells from the data as well as cells labeled with antibody hashtag #1 (Biolegend 394661) in batch 3, which were used solely as carrier cells for the 10X reaction. Clusters were defined with the resolution parameter set to 0.5. Differentially expressed genes between clusters were identified using the Seurat FindAllMarkers function with default parameters. Differentially expressed genes for 11 resulting clusters can be found in Extended data Table 3. R scripts for the final Seurat object generation can be found on GitHub (https://github.com/pogorely/COVID_vax_CD8).

### Donor and epitope assignment using feature barcodes

Cells were processed in 6 batches with each batch making a separate 10X Chromium reaction. In each batch, individual PBMC samples were uniquely labeled with a combination of DNA-barcoded hashing antibody (TotalSeq-C anti-human Hashtag antibodies 1-10, Biolegend) and a set of DNA-barcoded MHC-multimers. We attributed a cell to a certain hashtag if more than 50% of UMIs derived from hashing antibodies matched that hashtag. Cells specific to certain dextramers were called similarly: we required more than 30% of dextramer-derived UMIs to contain a dextramer-specific barcode, and if multiple dextramers passed this threshold the cell was considered specific to both. If the most abundant dextramer barcode per cell was ≤ 3 UMIs, we did not assign any epitope specificity to it. Cells were assigned to donors using a combination of hashing antibody and dextramer barcode. TCRα and TCRβ sequences were assembled from aggregated VDJ-enriched libraries using the CellRanger (v. 6.0.0) vdj pipeline. For each cell we assigned the TCRβ and TCRα chain with the largest UMI count. The R script performing feature barcode deconvolution, GEX and TCR join is available on Github (https://github.com/pogorely/COVID_vax_CD8) as well as the resulting Extended data Table 4.

### TCR repertoire analysis

T cell clones were defined as groups of cells from the same donor with identical nucleotide sequences of both CDR3α and CDR3β (see Extended data Table 5 for unique T cell clones). To correct erroneous or missing dextramer assignments for individual cells within a clone we assign each T cell a specificity of the majority of cells from this clone. To measure the distance between TCR α/β clonotypes and plot logos for dominant motifs we used the TCRdist algorithm implementation and plotting functions from *conga* python package^58^. Sequence similarity network analysis and visualizations were performed with the *igraph* R package^59^ and *gephi* software^60^. We exclude top 1% of vertices and edges with largest betweenness centrality values (which are likely to occur due to cell doublets or artifacts of scTCR sequencing) to filter out a small number of spurious connections between motif clusters A TCR motif cluster is then defined as a connected component on a similarity network. TCRβ repertoire diversity calculation was performed using normalized Shannon entropy 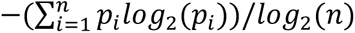, where *n* is a total number of unique TCRβ clonotypes, and *pi* is a frequency of *i-th* TCRβ clonotype (defined as the fraction of cells with this TCRβ amongst all cells in a sample with defined TCRβ).

### Artificial antigen-presenting cells (aAPCs)

A gBlock gene fragment encoding full-length HLA-A*01:01, HLA-A*02:01, HLA*A24:02 and HLA-B*15:01 was synthesized by Genscript and cloned into the pLVX-EF1α-IRES-Puro lentiviral expression vector (Clontech). Lentivirus was generated by transfecting HEK 293T cells (American Type Culture Collection (ATCC) CRL-3216) with the pLVX lentiviral vector containing the HLA insert, psPAX2 packaging plasmid (Addgene plasmid #12260), and pMD2.G envelope plasmid (Addgene plasmid #12259). Viral supernatant was harvested and filtered through a 0.45 µm SFCA syringe filter (Thermo Fisher) 24- and 48-hours post-transfection, then concentrated using Lenti-X Concentrator (Clontech). K562 cells (ATCC CCL-243) were transduced, then antibiotic selected for one week using 2 µg/mL puromycin in Iscove’s Modified Dulbecco’s Medium (IMDM; Gibco) containing 10% FBS and 1% penicillin/streptomycin. Surface expression of HLA was confirmed via flow cytometry using antibodies against HLA-A, B, C (PE-conjugated, Biolegend 311406, clone W6/32).

### TCR-expressing Jurkat 76.7 cells

TCR chains matching both the biggest clusters of Fig 4B, as well as the B15_NQK-specific prediction from^52^, were selected for Jurkat cell line generation (Extended data Table 6). TCRα and TCRβ chains for the selected epitope-specific TCRs were modified to use murine constant regions (murine TRAC*01 and murine TRBC2*01). A gBlock gene fragment was synthesized by Genscript to encode the modified TCRα chain, the modified TCRβ chain, and mCherry, with all three genes linked together by 2A sites. This sequence was cloned into the pLVX-EF1α-IRES-Puro lentiviral expression vector (Clontech). Lentivirus was generated by transfecting HEK 293T cells (ATCC CRL-3216) with the pLVX lentiviral vector containing the TCR-mCherry insert, psPAX2 packaging plasmid (Addgene plasmid #12260), and the pMD2.G envelope plasmid (Addgene plasmid #12259). Viral supernatant was harvested and filtered through a 0.45 µm SFCA syringe filter (Thermo Fisher) 24- and 48-hours post-transfection, then concentrated using Lenti-X Concentrator (Clontech). Jurkat 76.7 cells (a gift from Wouter Scheper; variant of TCR-null Jurkat 76.7 cells that expresses human CD8 and an NFAT-GFP reporter) were transduced, then antibiotic selected for 1 week using 1 µg/mL puromycin in RPMI (Gibco) containing 10% FBS and 1% penicillin/streptomycin. Transduction was confirmed by expression of mCherry, and surface TCR expression was confirmed via flow cytometry using antibodies against mouse TCRβ constant region (PE-conjugated, Biolegend 109208, clone H57-597) and human CD3 (Brilliant Violet 785-conjugated, Biolegend 344842, clone SK7).

### Intracellular cytokine staining functional assay

Jurkat 76.7 cells expressing the B15_NQK-specific TCR (2.5×10^5^) were co-cultured with HLA-B*15:01 aAPCs (2.5×10^5^) pulsed with 1 µM of either NQKLIANAF peptide from HKU1/OC43 common cold coronaviruses or NQKLIANQF peptide from SARS-CoV-2, 1 µg/mL each of anti-human CD28 (BD Biosciences 555725) and CD49d (BD Biosciences 555501), brefeldin A (GolgiPlug, 1 µL/mL; BD Biosciences 555029), and monensin (GolgiStop, 0.67 µL/mL; BD Biosciences 554724). An unstimulated (CD28, CD49d, brefeldin A, monensin) and positive control (brefeldin A, monensin, 1X Cell Stimulation Cocktail, PMA/ionomycin; eBioscience 00-4970-93) were included in each assay. Cells were incubated for 6 hours (37 °C, 5% CO-_2_), washed twice with FACS buffer (PBS, 2% FBS, 1 mM EDTA), then blocked using human Fc-block (BD Biosciences 564220) for 10 minutes at room temperature. The blocked cells were then stained with 1 µL Ghost Dye Violet 510 Viability Dye (Tonbo Biosciences 13-0870-T100) and a cocktail of surface antibodies 1 µL each of anti-human CD8 (Brilliant Violet 785-conjugated, Biolegend 344740, clone SK1), anti-human CD3 (Brilliant Violet 421-conjugated, Biolegend 344834, clone SK7), and anti-mouse TCRβ chain (PE-conjugated, Biolegend 109208) or APC/Fire750-conjugated, Biolegend 109246), clone H57-597) for 20 minutes at room temperature. Surface-stained cells were washed twice with FACS buffer, then fixed and permeabilized using the Cytofix/Cytoperm Fixation/Permeabilization kit (BD Biosciences) according to the manufacturer’s instructions. Following fixation and permeabilization, cells were washed twice with 1X Perm/Wash buffer and then stained with a cocktail of intracellular antibodies including 1.25 µL of anti-human IFNγ (Alexa Fluor 647-conjugated, Biolegend 502516, clone 4S.B3) and 1µL anti-human CD69 (PerCP-eFluor710-conjugated, eBioscience 46-0699-42, clone FN50) at 4 °C for 30 minutes. Cells were washed twice with 1X Perm/Wash buffer, and then were analyzed by flow cytometry on a custom-configured BD Fortessa using FACSDiva software (Becton Dickinson). Flow cytometry data were analyzed using FlowJo v. 10.7.1 software (TreeStar). Responsiveness to peptide stimulation was determined by measuring frequency of NFAT-GFP, IFNγ, and CD69 expression.

### Specificity validation of generated Jurkat cell lines

Jurkat 76.7 cells expressing the epitope-specific TCRs (1.5×10^5^) were co-cultured with aAPCs (1.5×10^5^) expressing the corresponding restricting HLA allele, and pulsed with 1 µM of cognate SARS-CoV-2 peptide, 1 µg/mL each of anti-human CD28 (BD Biosciences 555725) and CD49d (BD Biosciences 555501). An unstimulated (CD28, CD49d) and positive control (1X Cell Stimulation Cocktail, PMA/ionomycin; eBioscience 00-4970-93) were included for each Jurkat 76.7 cell line. Cells were incubated for 8 hours (37 °C, 5% CO-_2_) then washed with FACS buffer (PBS, 2% FBS, 1 mM EDTA), resuspended in 50µL FACS buffer, and blocked using human Fc-block (BD Biosciences 564220) for 10 minutes at room temperature. Cells were then stained with 1 µL Ghost Dye Violet 510 Viability Dye (Tonbo Biosciences 13-0870-T100) and a cocktail of surface antibodies including 1 µL each of anti-human CD3 (Brilliant Violet 421-conjugated, Biolegend 344834, clone SK7), 1 µL anti-human CD69 (PerCP-eFluor710-conjugated, eBioscience 46-0699-42, clone FN50), and anti-mouse TCRβ chain (APC/Fire750-conjugated (Biolegend 109246), clone H57-597). Cells were incubated for 20 minutes at room temperature and then washed with FACS buffer. Cells were analyzed by flow cytometry on a custom-configured BD Fortessa using FACSDiva software (Becton Dickinson). Flow cytometry data were analyzed using FlowJo software version 10.7.1 (TreeStar). Responsiveness to peptide stimulation was determined by measuring frequency of NFAT-GFP and CD69 expression.

To further test the specificity of generated Jurkat T cell lines we used dextramer staining with the same dextramer reagents used for staining PBMCs (above). Jurkat cells were washed with FACS buffer and resuspended in 50 µL. Cells were blocked with using human Fc-block (BD Biosciences 564220) and then stained with 1 µL of corresponding dextramer and 1 µL Ghost Dye Violet 510 Viability Dye (Tonbo Biosciences 13-0870-T100). A control Jurkat Cell line with known irrelevant specificity was used as a negative control and was stained with all dextramer reagents tested. All cells were stained for 40 minutes on ice. After the incubation cells were washed once with FACS buffer. Cells were analyzed by flow cytometry on a custom-configured BD Fortessa using FACSDiva software (Becton Dickinson). Flow cytometry data were analyzed using FlowJo software version 10.7.1 (TreeStar).

### Tetramer generation and staining of cross-reactive Jurkat Cell line

Biotinylated HLA-B*15-monomers loaded with NQKLIANQF (SARS-CoV-2) and NQKLIANAF (CCCoV) versions of the peptide were tetramerised using TotalSeq-C-0951-PE-Streptavidin (Biolegend 405261, 0.5 mg/mL) and TotalSeq-C-0956-APC-Streptavidin (Biolegend 405283, 0.5 mg/mL). 60 µL of HLA-monomers (500 nM) were mixed with 1 µL of PE-conjugated (B15_NQKLIANQF) or APC-conjugated (B15_NQKLIANAF) streptavidin reagents and incubated for 1 hour in the dark on ice. Jurkat 76.7 cells expressing the potentially cross-reactive TCR were stained with 1 µL Ghost Dye Violet 510 Viability Dye (Tonbo Biosciences 13-0870-T100) and 5 µL of each MHC-tetramer for 30 minutes on ice. Flow cytometry data were analyzed using FlowJo software (TreeStar). Cross-reactivity of the Jurkat 76.7 T cell line was determined by co-staining of the live cells with PE and APC-labeled MHC-tetramers.

### Recombinant SARS-CoV-2 proteins and ELISA

Expression plasmids for the nucleocapsid (N) protein, spike protein, and the spike receptor binding domain (RBD) from the Wuhan-Hu-1 isolate were obtained from Florian Krammer (Icahn School of Medicine at Mount Sinai). Proteins were transfected into Expi293F cells using a ExpiFectamine 293 transfection kit (Thermo Fisher Scientific) as previously described^61^. Supernatants from transfected cells were harvested and purified with a Ni-NTA column.

For hCoV and SARS-CoV-2 antibody detection, 384-well microtiter plates were coated overnight at 4 °C, with recombinant proteins diluted in PBS. Optimal concentrations for each protein and isotype were empirically determined to optimize sensitivity and specificity. SARS-CoV-2 spike RBD was coated at 2 µg/mL in PBS. Full-length spike was coated at 2 µg/mL for IgG. N protein was coated at 1 µg/mL. The spike proteins of hCoV-229E (Sino Biological, 40605-V08B), hCoV-NL63 (Sino Biological, 40604-V08B), hCoV-HKU1 (Sino Biological, 40606-V08B), or hCoV-OC43 (Sino Biological, 40607-V08B) were coated at 1 µg/mL for IgG detection. For all ELISAs, plates were washed the next day three times with 0.1% PBS-T (0.1% Tween-20) and blocked with 3% Omniblok^TM^ non-fat milk (AmericanBio; AB10109-01000) in PBS-T for one hour. Plates were then washed and incubated with plasma samples diluted 1:50 in 1% milk in PBS-T for 90 minutes at room temperature. Prior to dilution, plasma samples were incubated at 56 °C for 15 minutes. ELISA plates were washed and incubated for 30 minutes at room temperature with anti-human secondary antibodies diluted in 1% milk in PBS-T: anti-IgG (1:10,000; Invitrogen, A18805). The plates were washed and incubated at room temperature with OPD (Sigma-Alrich, P8287) for 10 minutes (for hCoV ELISAs) or SIGMAFAST OPD (Sigma-Alrich; P9187) for 8 minutes (for SARS-CoV-2 ELISAs). The chemiluminescence reaction was stopped by addition of 3N HCl and absorbances were measured at 490 nm on a microplate reader. The OD of each sample was normalized to the OD of the same two positive control samples that were run on each plate. The normalized OD is the percent ratio of the sample OD to the average OD of the positive controls for the plate. For the SARS-CoV-2 ELISAs, we first screened samples from prior studies that were collected before 2019 to identify the background level of the assay. Samples were considered positive if the normalized OD was greater than two times the average of normalized ODs from all SARS-CoV-2 negative samples in the SJTRC cohort (n=912). For the hCoV ELISAs, we screened samples from a prior study that included very young children to identify samples to serve as negative controls. Samples with a normalized OD greater than three times the average of the normalized ODs for the negative controls were considered positive for the hCoV antigens. Antibody levels for each donor can be found in the Extended data Table 2.

### Analysis of epitope mutations in SARS-CoV-2 variants

We used the WHO definition of variant of concern and variant of interest updated January 10, 2022. A mutation was included in the analysis if it appears in at least 10% of the GISAID (www.gisaid.org/hcov19-variants/, accessed on Dec 7 2021) isolates with the same Pango lineage and appears in >1000 isolates from that Pango lineage (Rambaut et al. 2020). To analyze the predicted binding of variant and wild type peptides, we used NetMHCpan 4.1b^47^. Results of this analysis are in Extended data Table 7.

### Statistical analysis

Statistical analysis was performed in R version 4.0.2. Wilcoxon signed-rank test was used to compare paired pre-vaccination and post-vaccination samples; only donors with cells collected at both timepoints were included in the test. Wilcoxon rank-sum test (Mann-Whitney U test) was used to compare unpaired samples between pairs of study groups, Kruskal-Wallis H test was used to test for difference between multiple study groups. Multiple testing correction was performed using the Benjamini-Hochberg procedure. Ns not significant, * p<0.5, **p<0.01, ***p<0.001

## Data Availability

Code required to reproduce source data for figures is available on GitHub: https://github.com/pogorely/COVID_vax_CD8. All data produced in the study is available as supplementary files. Raw sequencing data was deposited to Short Read Archive acc. PRJNA744851.

## Acknowledgements

We thank all the donors who volunteered for the SJTRC study, Phil Bradley and Stefan Schattgen for their consultations on TCRdist and conga algorithms, Greig Lennon from St. Jude Immunology flow core for his help with FACS. This work was funded by ALSAC at St. Jude, the Center for Influenza Vaccine Research for High-Risk Populations (CIVR-HRP) contract number 75N93019C00052 (S.S-C, P.G.T), the St. Jude Center of Excellence for Influenza Research and Surveillance (S.S-C, M.A.M, P.G.T), HHSN272201400006C, 3U01AI144616-02S1 (P.G.T, M.A.M, S.S-C), and R01AI136514 (P.G.T).

## Author Contributions

Conceptualization: A.A.M, M.V.P, E.K.A, J.C.C., P.G.T. Formal analysis: A.A.M, M.V.P, A.M.K, J.C.C, M.A.M, J.H.E, X.Z, K.V, G.W. Investigation: A.A.M., M.V.P, A.M.K, C-H.C, R.C.M, M.A.M, J.W, J.E., C-Y.L, D.B, S.T, P.K, D.D. S.M, S.R.O. Methods development: A.A.M, M.V.P, J.C.C., A.M.K, C-Y.L, S.S-C, M.A.M. Resources: S.S-C, M.A.M, P.T, J.H.E., J.W. Data and sample curation: J.W, J.H.E, E.K.A, J.C.C., K.J.A, SJTRC Study Team. Writing, original draft: A.A.M. and M.V.P. Writing, review, and editing: A.A.M, M.V.P, A.M.K, E.K.A, J.C.C, J.W, M.A.M, P.G.T. Visualization: A.A.M., A.M.K. Supervision: P.G.T. Funding Acquisition: P.G.T.

## Extended data

**Extended data Table 1.** SARS-CoV-2 derived CD8^+^ epitopes used for MHC-multimer generation.

**Extended data Table 2**. Study participant metadata.

**Extended data Table 3.** Differentially expressed genes for GEX clusters of epitope-specific CD8+ T cells.

**Extended data Table 4.** Epitope-specific CD8^+^ T cells GEX clusters, TCR and epitope specificity.

**Extended data Table 5.** Unique epitope-specific CD8^+^ αβTCR clonotypes.

**Extended data Table 6:** TCR amino acid sequences used for generation of TCR-expressing Jurkat cell lines

**Extended data Table 7.** Mutations in studied epitopes from SARS-CoV-2 variants.

---

**Extended data Fig. 1.**
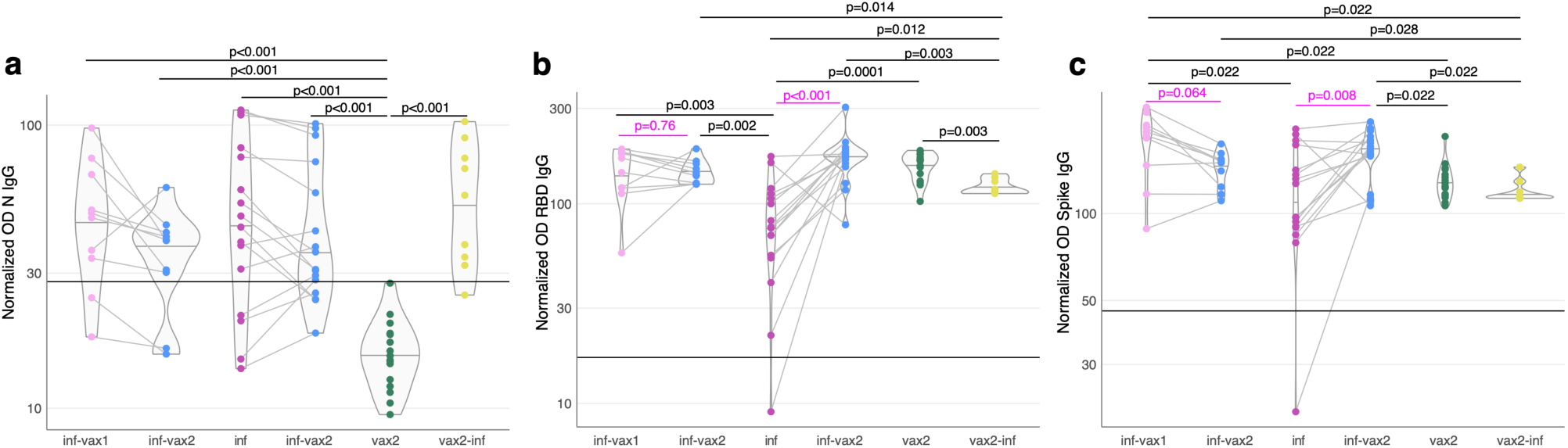
Antibody levels across study groups. Plasma was tested by ELISA for IgG antibodies specific for (**a**) Nucleocapsid (N), (**b**) the receptor-binding domain (RBD) of the spike, (**c**) whole spike protein of SARS-CoV-2. Normalized ODs are the percent ratio of the sample OD to the OD of the positive control samples for each plate. The black horizontal line on the plots indicates the positivity threshold, which is two times the average of the normalized ODs for all SARS-CoV-2 negative samples in the cohort. P-values for Mann-Whitney U test after Benjamini-Hochberg multiple testing correction are reported. Donors sampled before and after mRNA vaccination are connected with a line. P-values (magenta) for paired samples were calculated with the Wilcoxon signed-rank test.

**Extended data Fig. 2.**
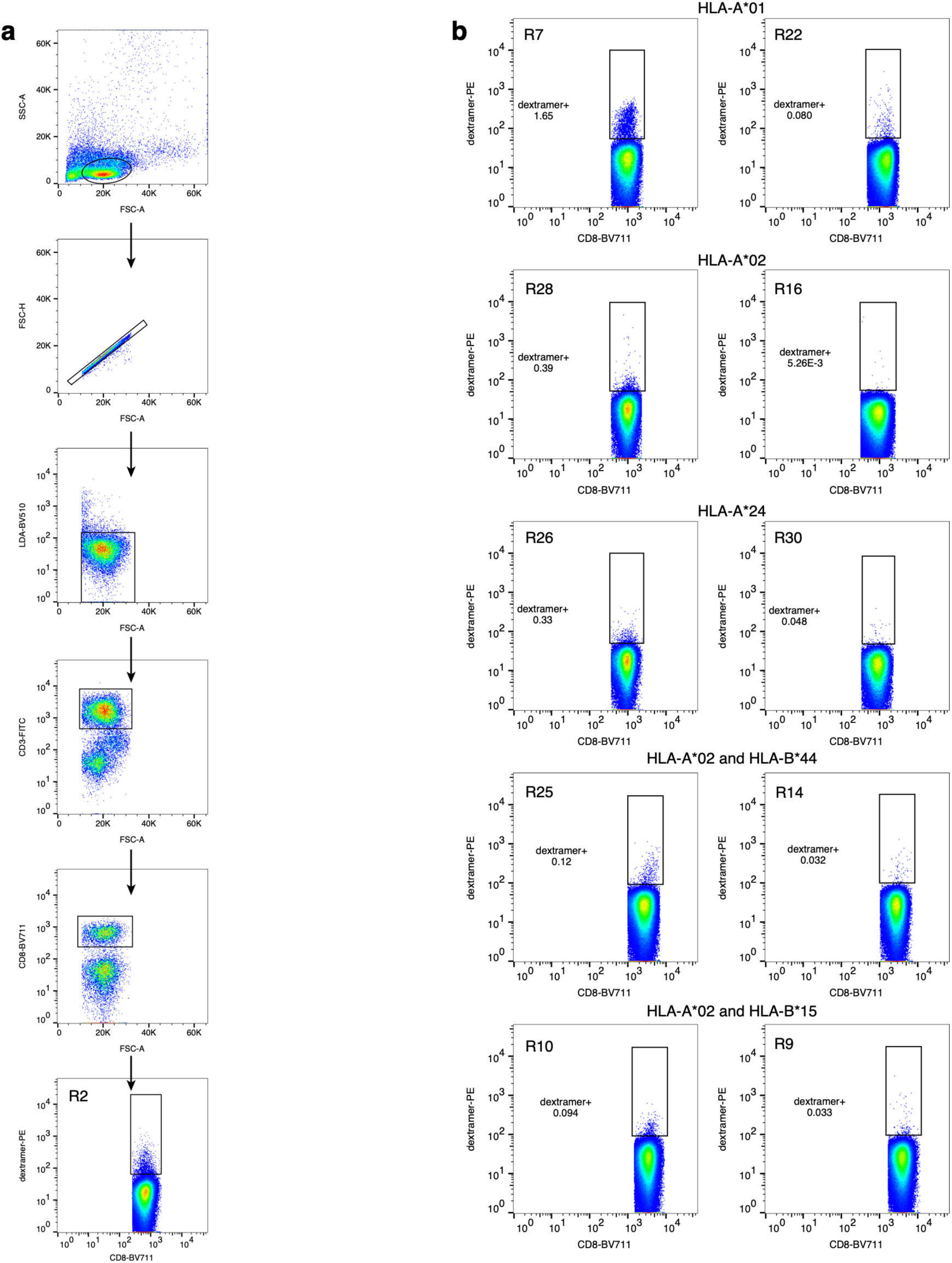
a. Gating strategy for sorting of single live CD3^+^CD8^+^dextramer^+^ cells. b. Representative flow plots for donors stained with the same dextramer pools, but showing different frequencies of single live CD3^+^CD8^+^dextramer^+^ cells.

**Extended data Fig. 3.**
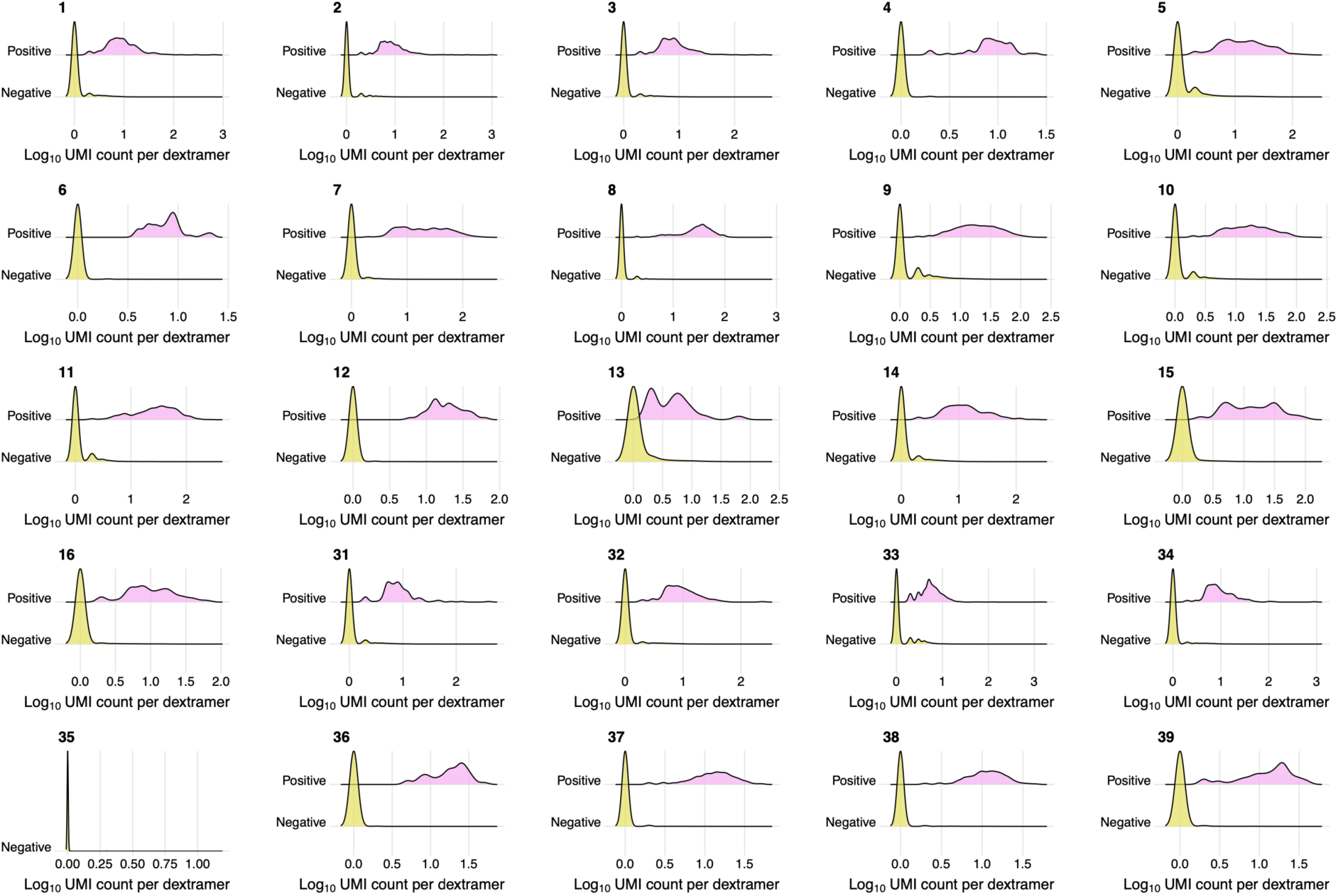
Dextramer assignment with feature barcodes. Each subplot shows distribution of Log_10_ (# UMIs) for dextramers with certain feature barcodes in dextramer-negative (yellow) and dextramer-positive (pink) cells. Dextramer with barcode 35 B44_VEN_M did not have any specific cells.

**Extended data Fig. 4.**
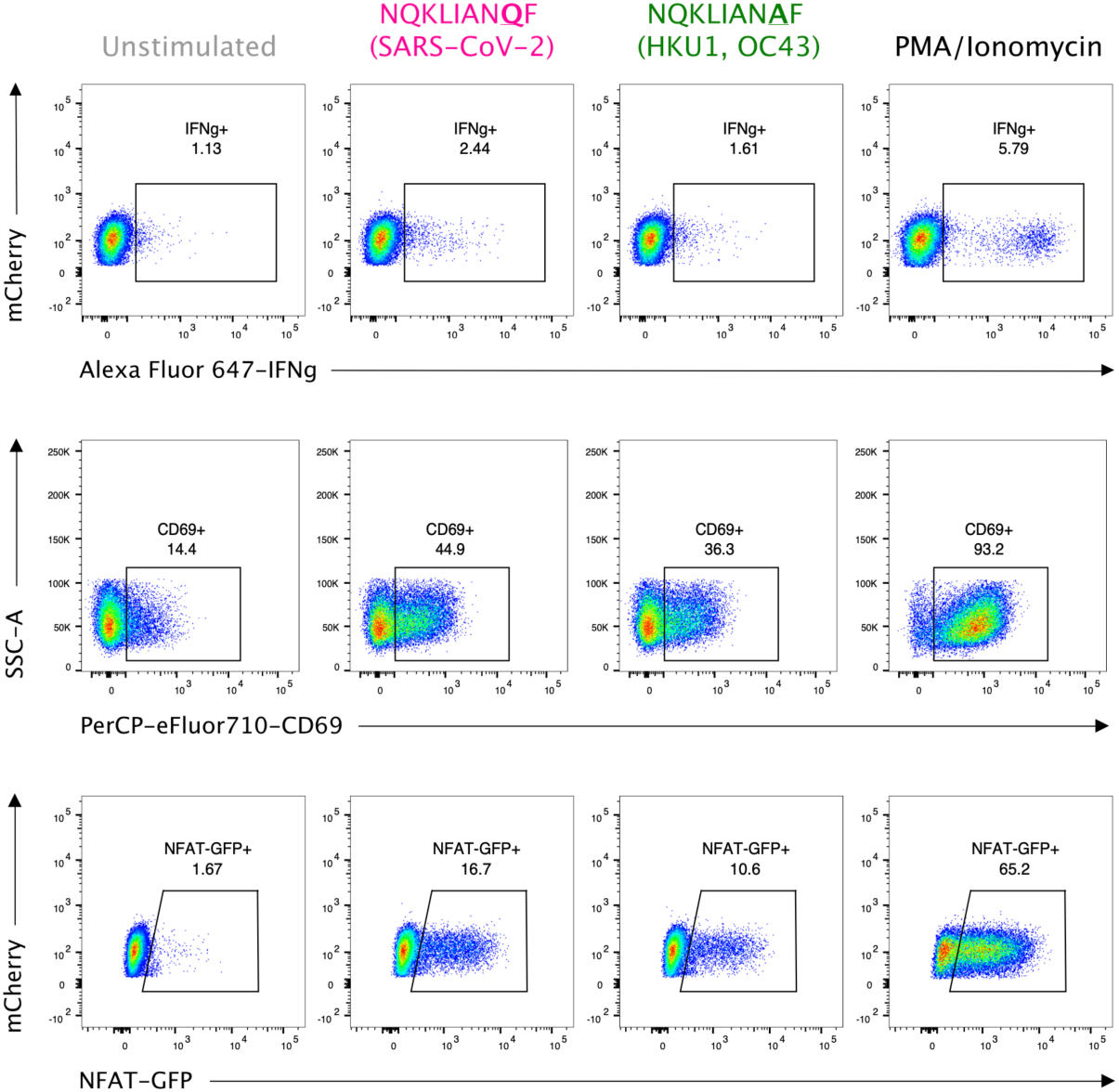
Peptide stimulation confirms cross-reactivity of B15_NQK αβTCR. From left to right: unstimulated (negative control), NQKLIANQF (SARS-CoV-2) peptide stimulation, NQKLIANAF (OC43 and HKU1) peptide stimulation, PMA/Ionomycin (positive control). Top row: IFN-γ production by TCR-expressing Jurkats measured by intracellular cytokine staining. Middle row: CD69+ surface expression. Bottom row: NFAT-GFP reporter expression.

**Extended data Fig. 5.**
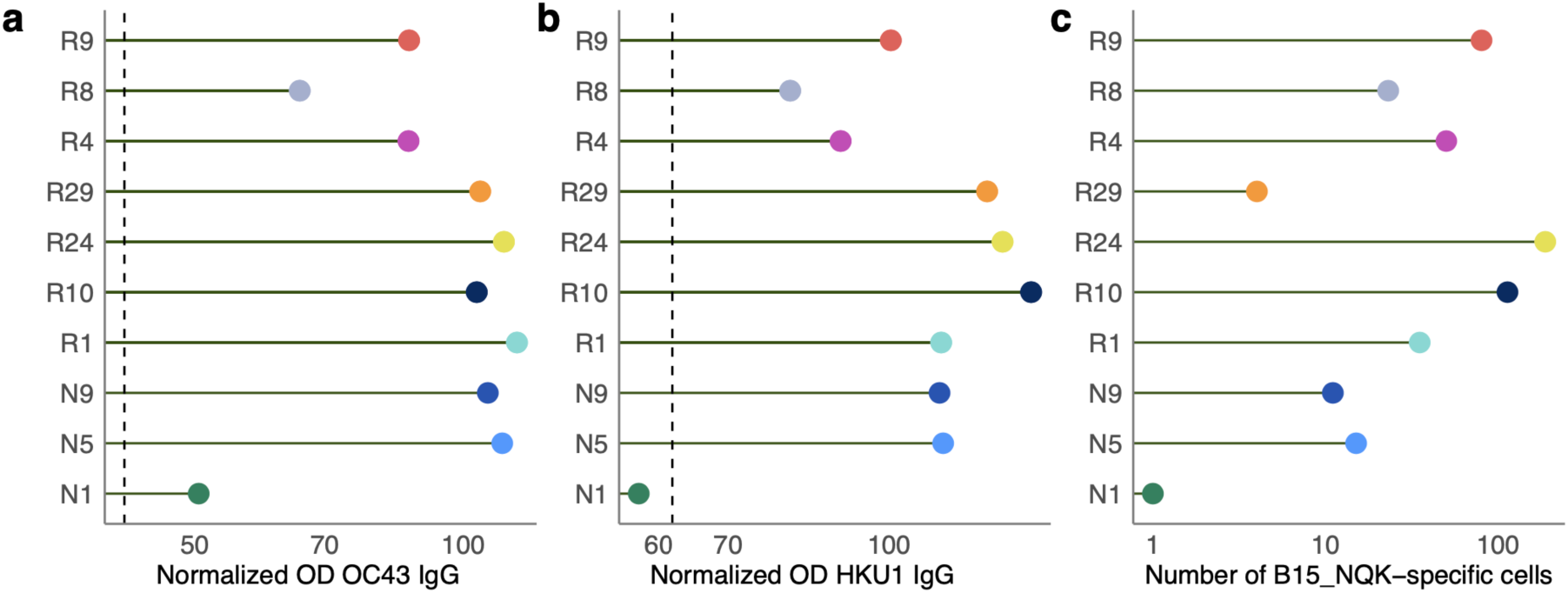
Antibody titers for CCCoV spike protein and number of B15_NQK cross-reactive cells in HLA-B*15:01^+^ donors. Plasma collected from donors prior to infection or vaccination was tested by ELISA for IgG antibodies to the spike of **a,** hCoV-OC43 or **b,** hCoV-HKU1. The normalized ODs are the percent ratio of the sample OD to the OD of the positive control sample for each plate. The dashed line is the threshold for positivity, which is three times the average of the normalized OD for the negative control samples. **c,** The number of HLA-B*15:01-restricted epitope T cells after infection or vaccination (log-scale).

**Extended data Fig. 6.**
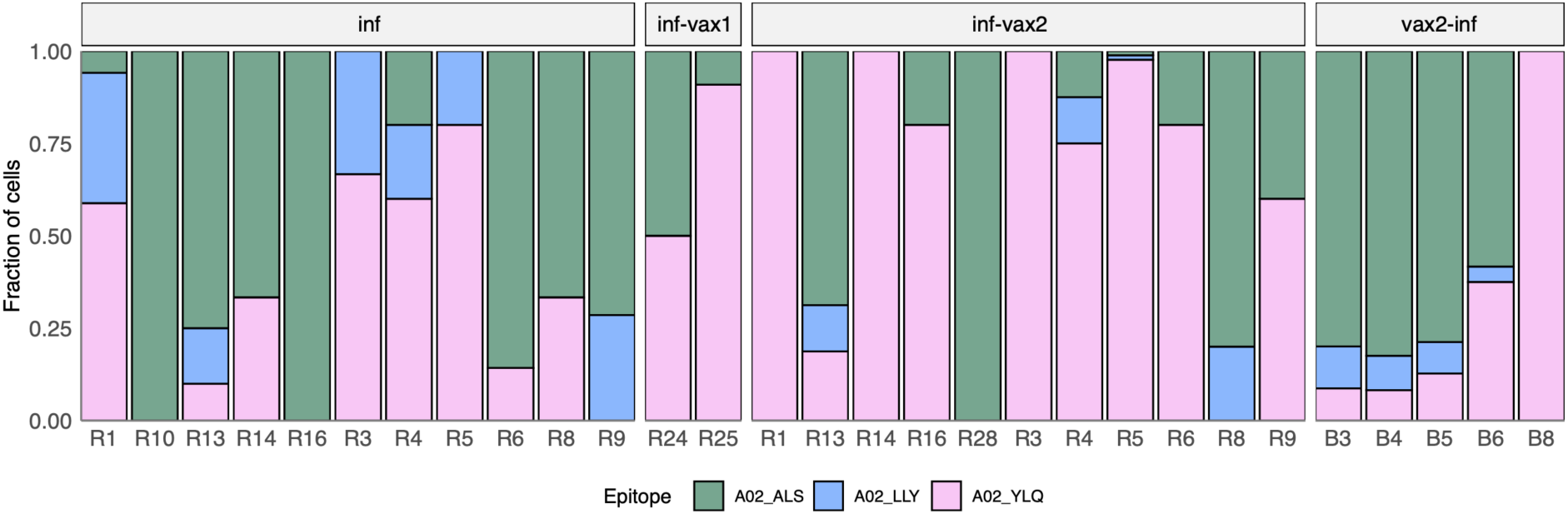
Composition of HLA-A*02-restricted T cell response in HLA-A*02 positive donors. Increasing proportion of spike-targeting T cells (pink) is observed after vaccination of previously infected individuals.

**Extended data Fig. 7.**
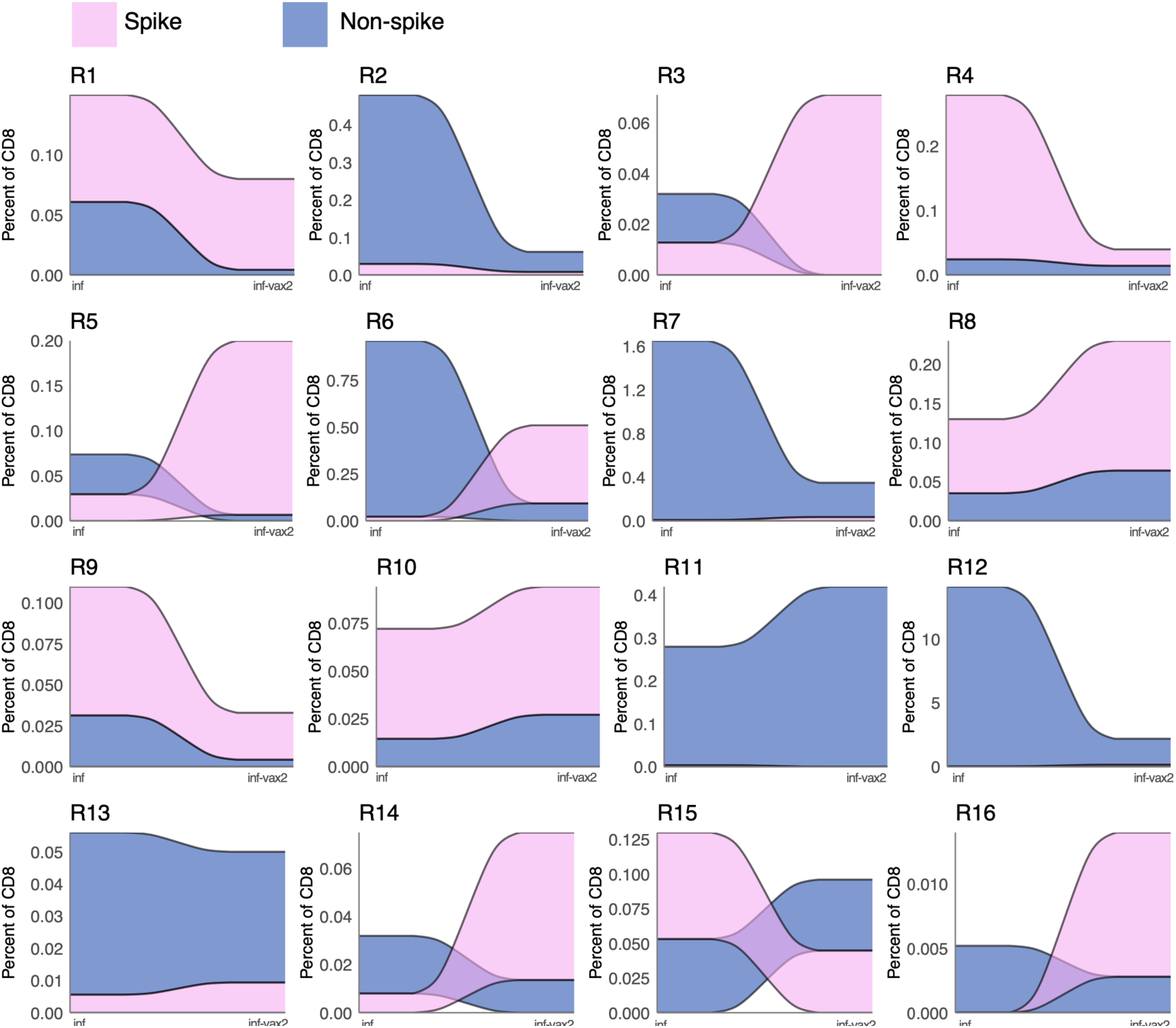
Clonal dynamics of spike- and non-spike-specific T cell response for SARS-CoV-2 infected donors before and after two doses of BNT162b2. Each colored ribbon represents an estimated frequency of spike- (pink) or non-spike- (blue) specific T cells.

**Extended data Fig. 8.**
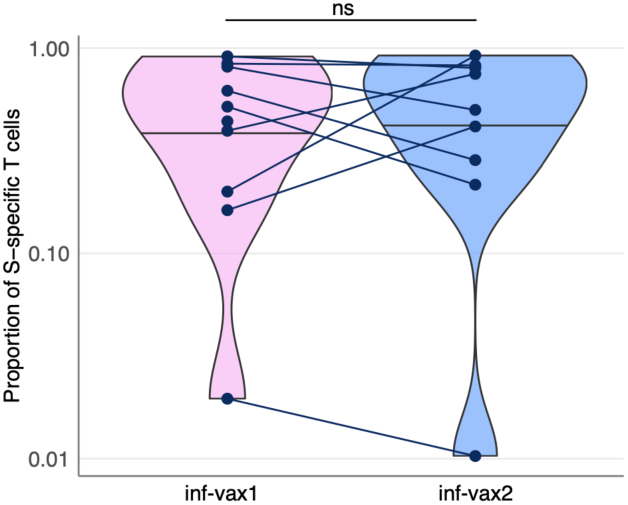
SARS-CoV-2 infected individuals after the first and second BNT162b2 vaccine doses (inf-vax1 and inf-vax2) have the same proportion of spike-specific T cells. (p=0.9, Wilcoxon signed-rank test). Spike T cell proportion was calculated as a fraction of spike-specific T cells out of all CD8^+^ epitope-specific T cells of a donor in scRNAseq data.

**Extended data Fig. 9.**
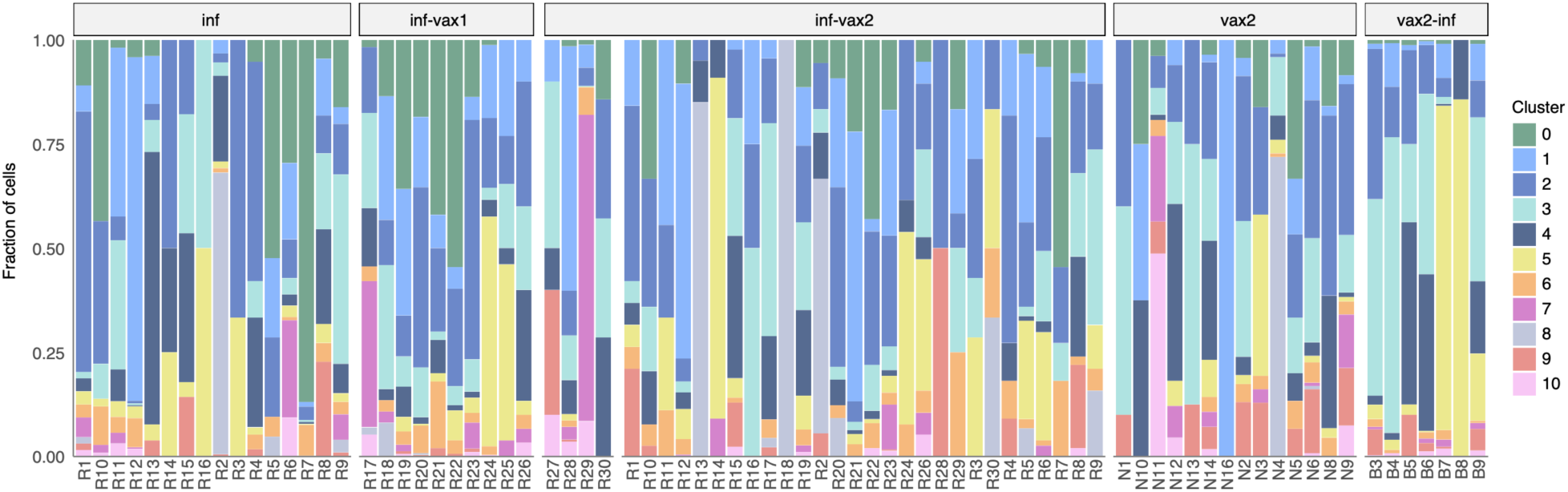
GEX cluster distribution for each sample. Each colored bar represents a fraction of cells in a given GEX cluster.

**Extended data Fig. 10.**
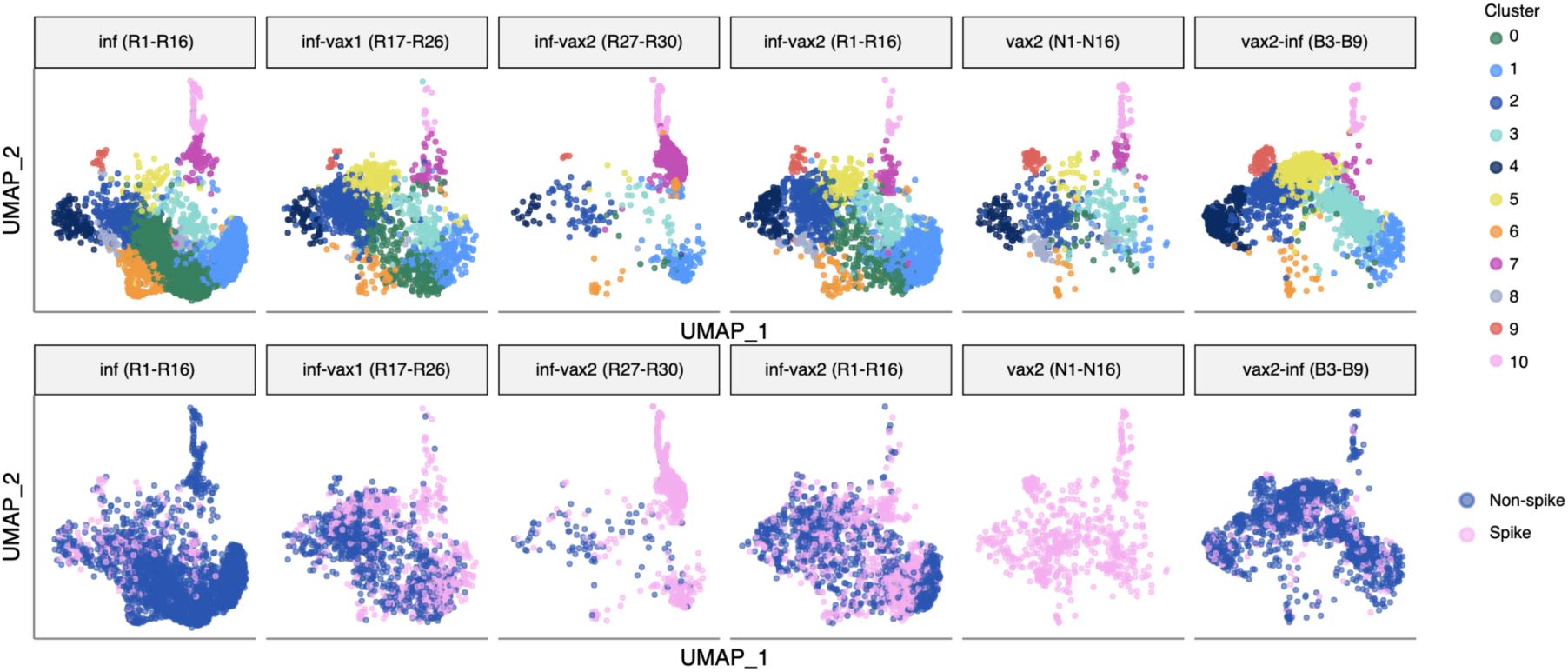
UMAP visualization of cells clustered by similarity of GEX. Each subpanel shows cells from each study group. Top: cells colored by cluster. Bottom: cells colored by spike and non-spike specificity.

**Extended data Fig. 11.**
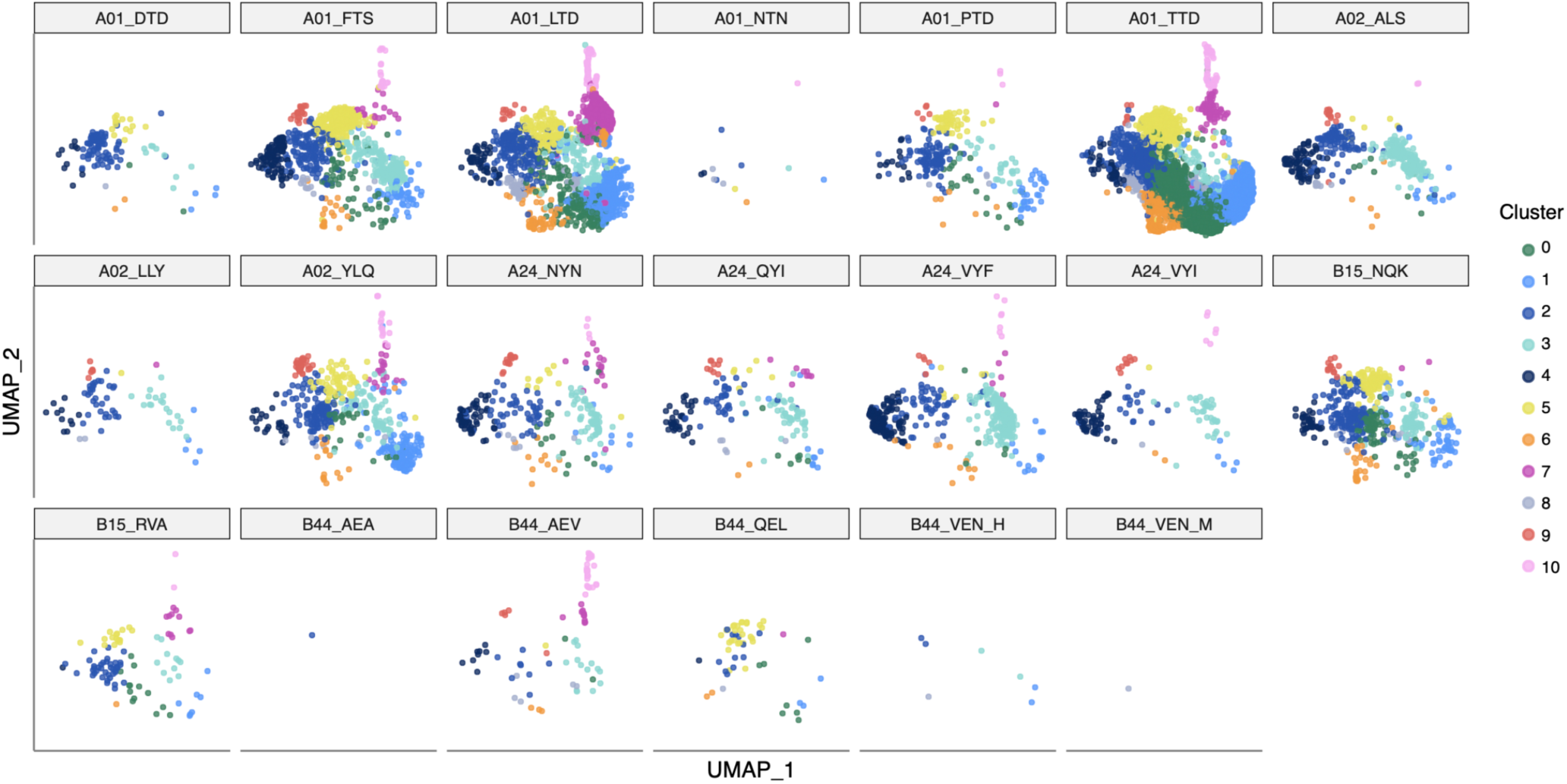
UMAP visualization of cells clustered by similarity of GEX. Each subpanel shows cells specific for each of the tested epitopes.

**Extended data Fig. 12.**
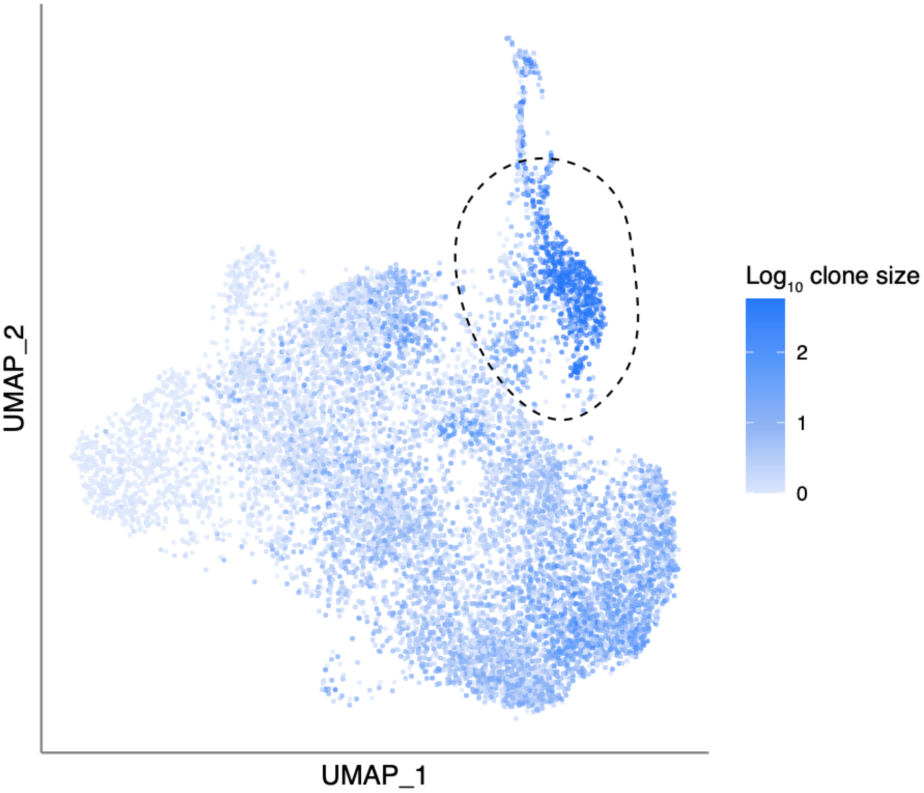
“Exhausted” cluster 7 (circled) is enriched with cells from expanded clones. The color of each dot shows the size of the T cell clone (Log_10_ of number of cells) for each cell.

**Extended data Fig. 13.**
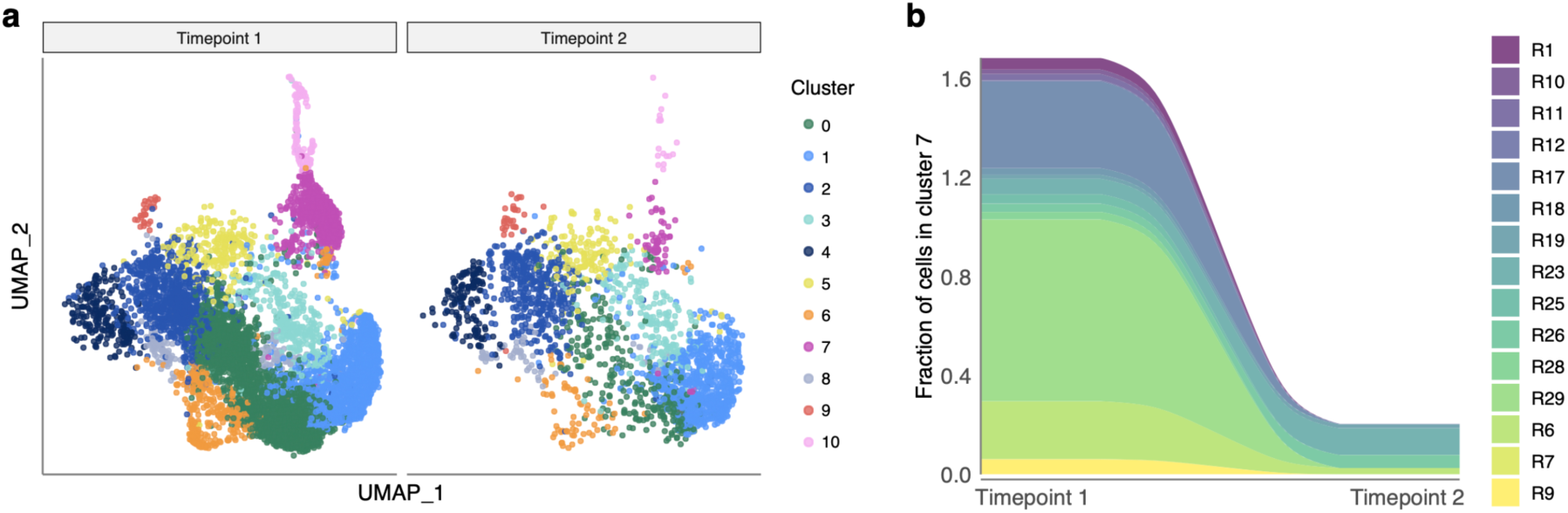
Number of cells in the “exhausted” cluster (cluster 7) declines over time. **a. UMAP visualization of cells clustered by similarity of GEX for donors sampled twice during the study (shapes connected with a line on** **Fig. 1b**). Timepoint 1 corresponds to inf (R1-R16), inf-vax (R17-R30); timepoint 2 corresponds to inf-vax2 (R1-R30). **b. Fraction of cells in cluster 7 out of all cells.** Only donors with cells in cluster 7 on timepoint 1 are shown.

**Extended data Fig. 14.**
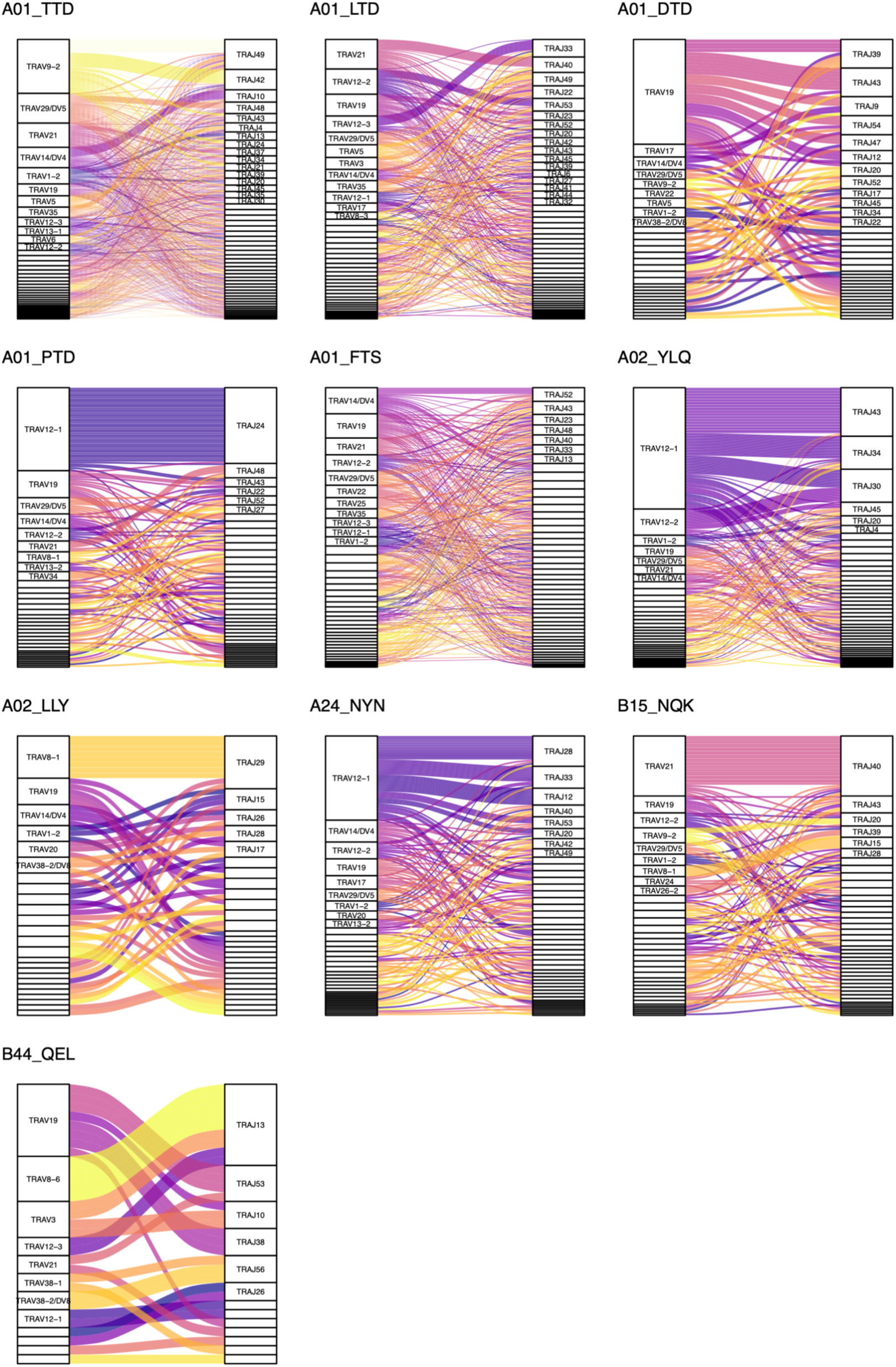
VαJα-usage for selected epitopes. Height of each rectangle corresponds to the fraction of unique epitope-specific T cell clones expressing a given V- or J-segment in the TCRα. Ribbons show the frequency of VJ combinations.

**Extended data Fig. 15.**
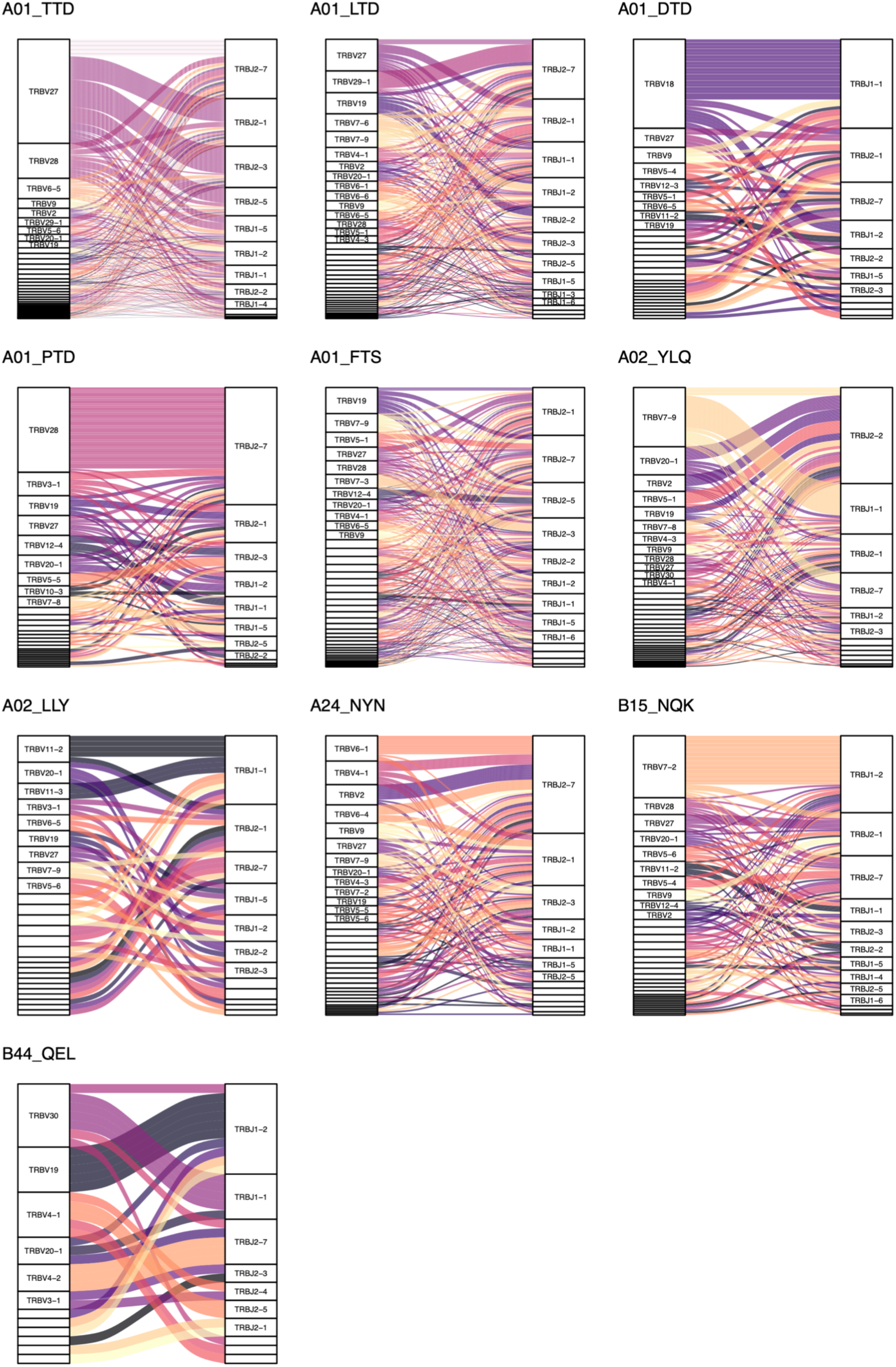
VβJβ-usage for selected epitopes. Height of each rectangle corresponds to the fraction of unique epitope-specific T cell clones expressing a given V- or J-segment in the TCRβ chain. Ribbons show the frequency of VJ combinations.

**Extended data Fig. 16.**
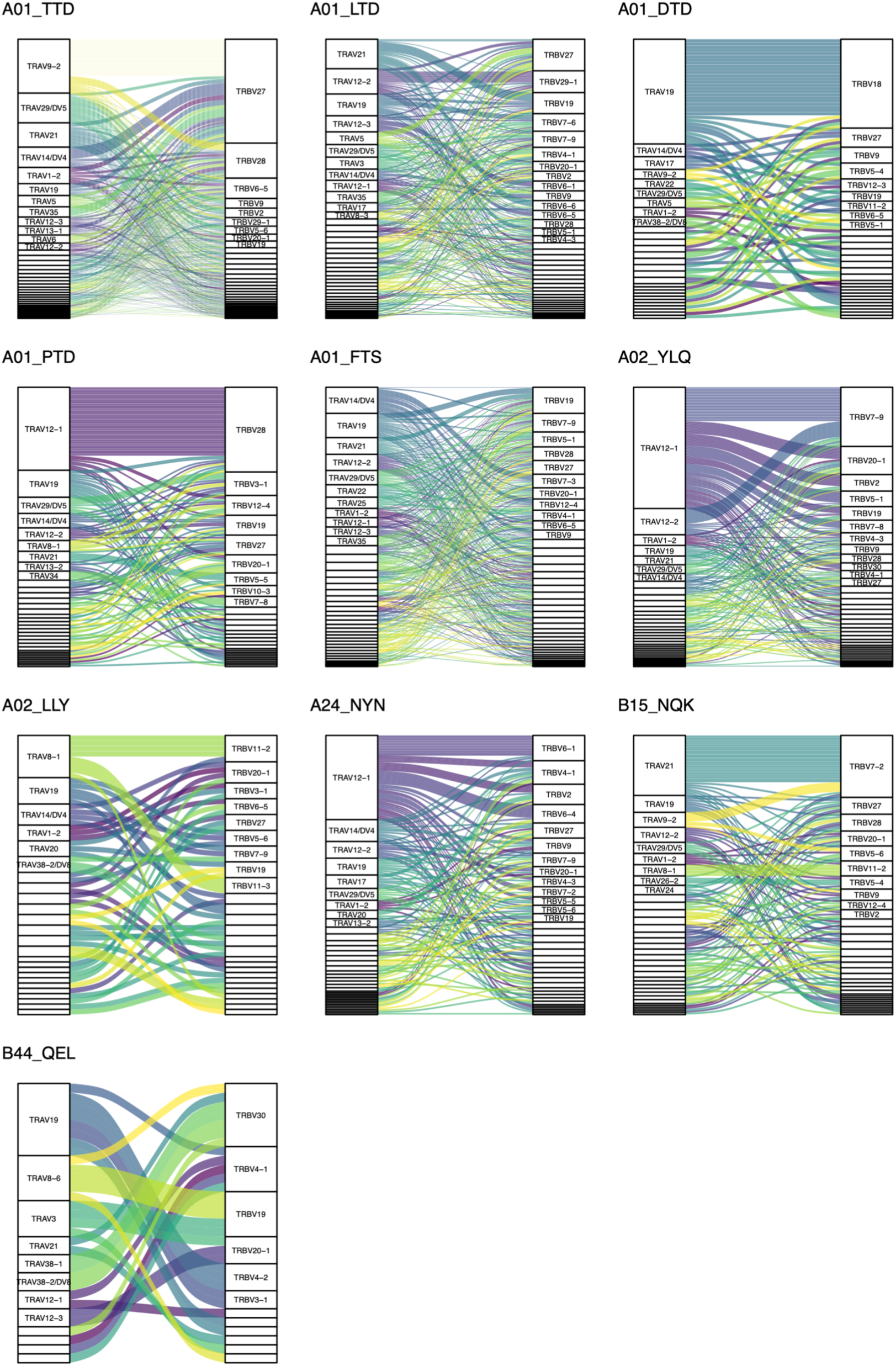
Vα-Vβ pairings for selected epitopes. Height of each rectangle corresponds to the fraction of unique epitope-specific T cell clones expressing a given TRAV or TRBV segment. Ribbons show frequencies of TRAV-TRBV combinations.

**Extended data Fig. 17.**
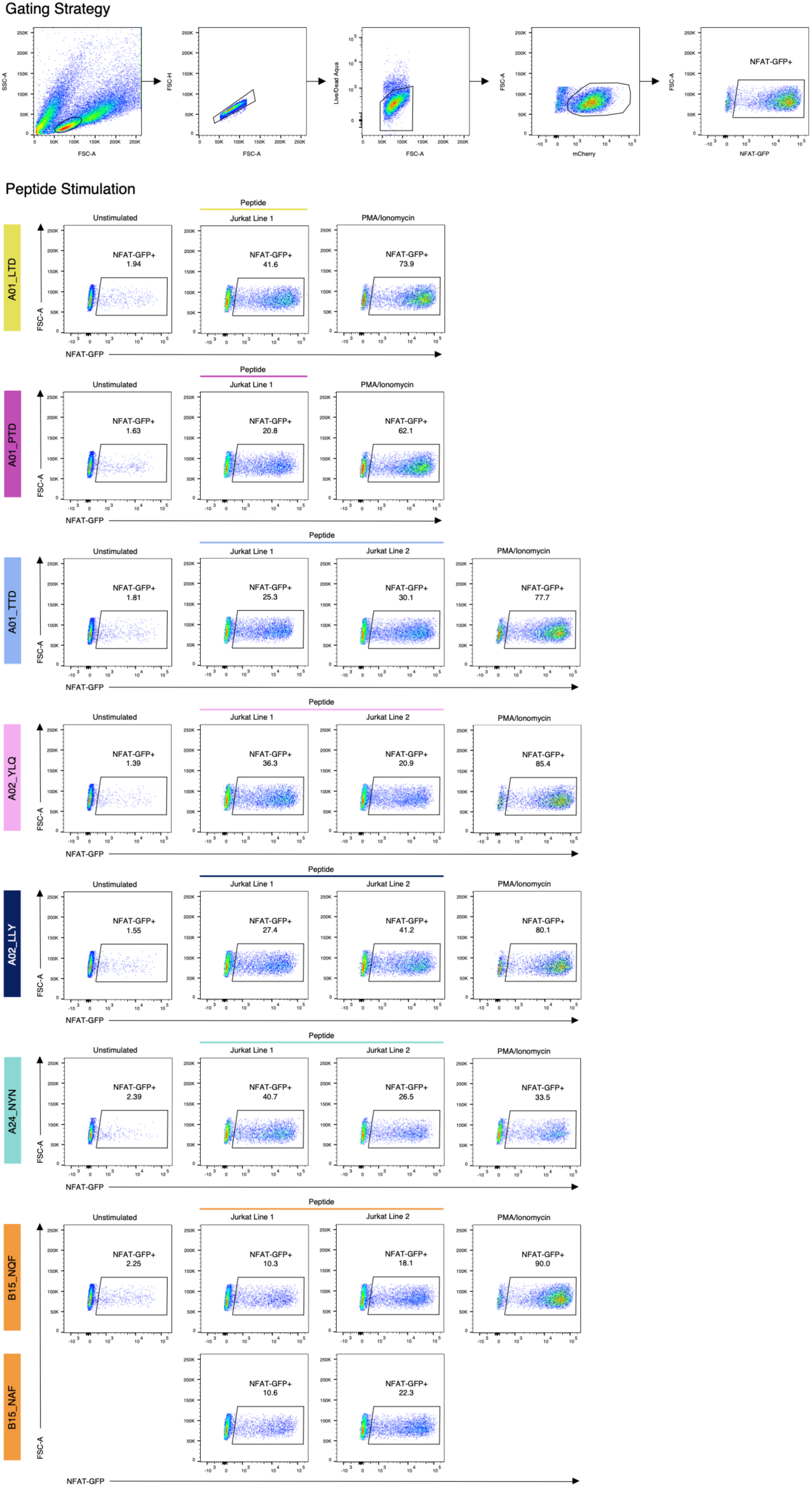
Peptide stimulation confirms specificity of αβTCR motifs. Top: example of the gating strategy (B15_specific Jurkat line 1, same as Fig. S4). Left column: unstimulated control. Each row shows stimulation with a single peptide (middle columns), B15 specific TCRs were stimulated with both NQKLIAN**Q**F (SARS-CoV-2) peptide and NQKLIAN**A**F (OC43 and HKU1) peptide; Right column: PMA/Ionomycin (positive control). Responsiveness of the Jurkat cell lines was determined using an endogenous NFAT-GFP reporter.

**Extended data Fig. 18.**
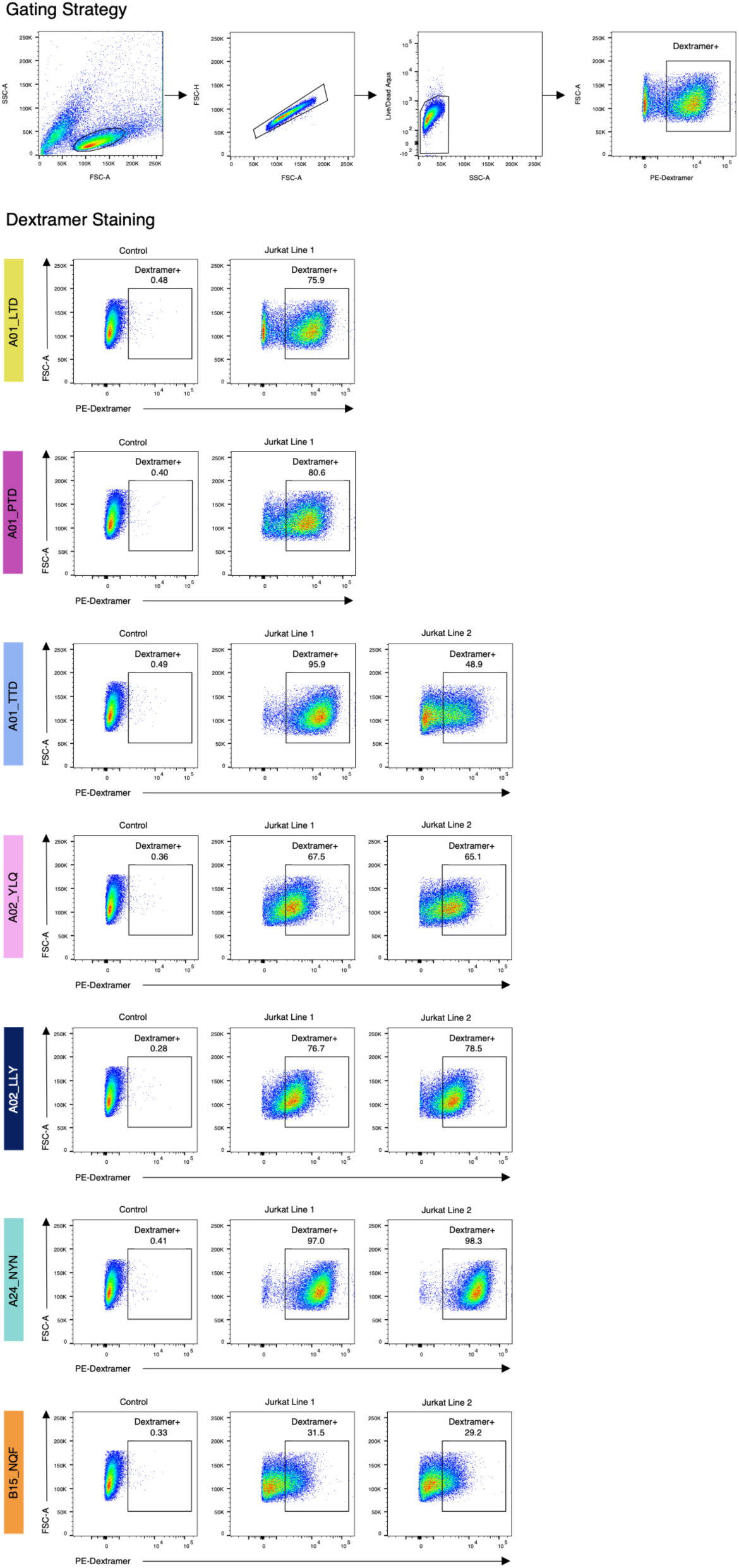
MHC-dextramer staining confirms specificity of αβTCR motifs. Top: example of the gating strategy (B15_specific Jurkat line 1, same as Fig. S4). Left column: control Jurkat cell line with other known specificity. Each row shows staining with a single MHC-dextramer.

**Extended data Fig. 19.**
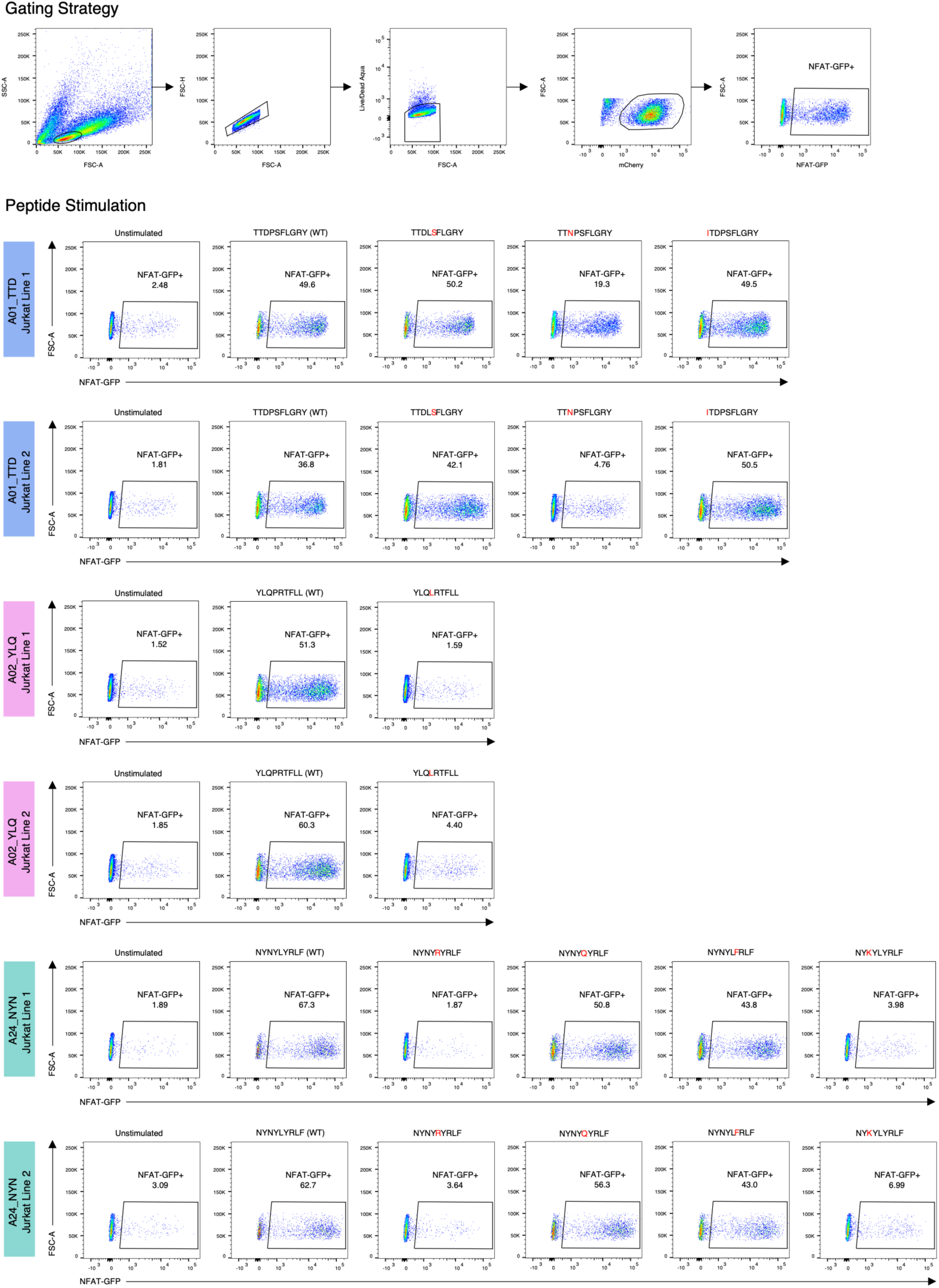
Recognition of SARS-CoV-2 mutated epitopes by αβTCR motifs. Left column: unstimulated control. Each row shows stimulation with a single peptide (middle columns). Responsiveness of the Jurkat cell lines was determined using an endogenous NFAT-GFP reporter.

